# Assessing adherence to the UK Government’s sugar, salt, and calorie reduction targets by the highest-grossing restaurants’ menus in 2024: A cross-sectional study

**DOI:** 10.1101/2025.08.01.25332693

**Authors:** Alice O’Hagan, Rachel Pechey, Hannah Forde, Lauren Bandy

## Abstract

**Background:** To address high rates of diet-related disease, the UK Government has a series of voluntary targets for retailers, manufacturers, and the out-of-home sector (e.g., restaurants), to reduce the sugar, salt, and calorie content of food products. There is limited evidence for whether the out-of-home sector is progressing towards these targets, and individual company responses have not been evaluated. This study aimed to assess adherence to UK Government’s sugar, salt, and calorie reduction targets for menu items offered by the highest-grossing restaurant chains in 2024.

**Methods and Findings:** Nutritional information was collected from restaurants’ online menus. Mean/median sugar, salt, and calorie content, per 100g and per serving, was calculated for each restaurant and food subcategory. Sugar, salt, and calorie content for each menu item was compared against the UK Government’s targets, and the proportion of menu items meeting (i) each and (ii) every applicable target, was calculated for each restaurant and food subcategory.

3099 menu items were included. Across all restaurants, 61% of menu items met their calorie targets, 58% met their salt targets, 36% met their sugar targets, and 43% met all of their applicable targets. Six of the 12 food subcategories, and nine of the 21 restaurants, had over 50% of menu items meeting all of their applicable targets. Papa John’s was the lowest adhering restaurant for the calorie (35%) and salt (8%) targets, and Burger King, KFC, Nando’s, and Vintage Inns for the sugar targets (0%). Restaurants offering predominantly Pizzas had the lowest adherence to all applicable targets (32% overall), but restaurants with similar menu foci also varied in their adherence, e.g., for the Sandwich restaurants, 76% of menu items from Subway met all of their applicable targets, compared to 30% from Caffé Nero.

**Conclusions:** Our findings suggest that whilst certain restaurant types appear to perform worse than others against the UK Government’s sugar, salt, and calorie targets, target adherence for restaurants with similar menu portfolios also varies, highlighting the potential for restaurants to improve the nutritional quality of their products without changing their menu focus. Our study demonstrated that voluntary schemes do not incite a high degree of engagement across the out-of-home sector, and therefore mandatory regulations may be a more effective approach to improving the nutritional quality of out-of-home food.

## Background

The purchasing and consumption of foods high in energy, saturated fat, free sugars, and salt, is associated with an increased risk of obesity and diet-related non-communicable diseases (NCDs) (1). Globally, diet-related NCD prevalence is high, with approximately 40% of cardiovascular disease mortality between 1990 and 2019 attributable to dietary risk factors (2). The same is true for the UK specifically, with poor diet being the second-leading cause of death and ill health (3).

Food reformulation initiatives are a common approach to NCD prevention across the globe; 68 countries have a salt reduction policy in place (4), and the UK, Brazil, New Zealand, and USA, have further initiatives for sugar reduction, for example (5). Modelling studies highlight the potential for such policies to significantly reduce the incidence of diet-related diseases, such as obesity and cardiovascular disease (6,7). The UK Government first introduced their salt reduction programme in 2004, following advice from the Scientific Advisory Committee on Nutrition to reduce the population average salt intake to 6g a day. The first reduction and reformulation targets for industry were published in 2006 and have since been revised on several occasions, with the most recent revision in 2020 covering 84 category-specific salt targets for grocery foods, and 24 for the out-of-home (OOH) sector (8). Evidence suggests that the salt reduction programme has been successful, with average urinary sodium reducing by approximately 2% each year from 2004 to 2011, and significant reductions being observed in the salt content of food products sold by UK retailers (9).

Complementary to the salt reduction programme, the UK Government published targets for sugar and calories in 2017 and 2020, respectively (10,11). The sugar reduction programme aimed to achieve a 20% reduction in the sugar content of 14 food categories that contribute most to sugar intake in children, across all sectors of the food industry, by 2020 (10). The calorie reduction programme aimed to achieve a 10% reduction by retailers and manufacturers across nine food categories, and a 20% reduction by the OOH sector across seven food categories (with four common categories), in the calorie content of products sold, by 2025 (extended from 2024 due to the Covid-19 pandemic) (11). Governmental progress reports indicate that within the retailer and manufacturer sector, there has been a 3.5% reduction in the sugar content of products sold between 2015 and 2020 (12), and category-level decreases of up to 2.4% in the calorie content of products sold between 2017 and 2021 (13). Their findings suggest comparatively less progress has been made in the OOH sector, with only a 0.2% reduction in the sugar content of products sold between 2017 and 2020, and category-level increases of up to 2.3% in the calorie content of products sold between 2017 and 2021 (12,13). However, since the most recent data included in the sugar, salt, and calorie reduction progress reports was from 2020, 2018, and 2021 respectively, these conclusions may have since changed.

A considerable proportion of UK individuals’ weekly food intake comes from takeaway or restaurant meals (14). Eating OOH is becoming more popular, with a 159% increase in per-person expenditure on eating out from 2021 to 2022 in the UK (15), and an additional 3.5% increase to 2023 (16), potentially due to its perceived lower cost and higher convenience than preparing food at home (17). The OOH sector is dominated by a small number of multinational companies who operate chained food outlets (hereafter ‘restaurants’), with some of the highest-grossing companies including McDonald’s, KFC, Domino’s, and Greggs. In 2023, sales from chained food service outlets reached £35.1 billion in the UK, increasing by nearly £8 billion over the previous two years (18), showcasing the significant influence these companies have over diet and health.

The UK Government’s sugar, salt, and calorie reduction targets are voluntary, leaving the responsibility for meeting the targets to individual companies, yet there is limited evidence for how food companies have responded to the reduction programmes. One study looking at company-level adherence to the sugar reduction targets among UK manufacturers found that of the top 10 companies across 5 target food categories, just under half met the interim 5% sugar reduction targets for 2018 (19). Similar work conducted in the OOH sector found that only four out of 48 brands reduced the sugar content of their desserts by at least 20% from 2018 to 2020, and only half of which significantly reduced their calorie content as well (20).

There is a need for a more comprehensive assessment of company-level adherence to the targets. Furthermore, fewer studies in the broader literature have assessed the nutritional quality of foods in the OOH sector compared to the retailer and manufacturer sector, due to OOH nutrition data being less easily accessible. Evaluating industry’s response to the reduction programmes will improve transparency around companies’ commitment to population health, help with enforcement of comparable schemes in the future, and underpin other approaches to incentivising healthier food provision (e.g., investment decision-making based on food healthiness). This study aimed to assess adherence to the UK Government’s sugar, salt, and calorie reduction targets for food items offered by the highest grossing restaurant chains in the UK, in 2024.

## Methods

We assessed the nutritional content of food items offered in 2024 by the 21 highest-grossing restaurant chains in the UK. We collected nutrition information for restaurants’ menu items directly from their websites. Each menu item was categorised by the sugar, salt and calorie reduction target groups and their nutrient values compared to the relevant targets. The average kcal, salt, sugar per 100g and per serving, was also calculated for each restaurant and food subcategory. The study protocol was published on Open Science Framework prior to data analysis (21). This study is reported as per the Strengthening the Reporting of Observational Studies in Epidemiology (STROBE) guideline (S1 STROBE checklist).

### Identifying Restaurants

We identified the 21 highest-grossing chained restaurants in the UK using the latest (2022) sales data from Euromonitor International, a privately-owned market research company.

Euromonitor’s data portal Passport GMID was accessed via the Bodleian Library, University of Oxford (22). We selected restaurants that Euromonitor defined as “Consumer Foodservice”, including “cafés/bars, full-service restaurants, limited-service restaurants, self-service cafeterias and street stalls/kiosks”. “Full-service” refers to sit-down restaurants with table service, whilst “limited-service” refers to fast food or take-away outlets.

We excluded restaurants that had separate menus for each individual site to avoid introducing considerable heterogeneity to nutritional information at the restaurant level. Restaurants that only provided nutrition information per ingredient (e.g., bun, patty, cheese) rather than per item (e.g., complete burger) were also excluded, as it was unclear what a full menu item would consist of.

We initially aimed for a sample of 20 restaurants. Subway was excluded at the start of the data collection phase as it provided nutrition information for individual ingredients only.

However, new menu-item nutrition information was subsequently published after our data collection had started, and therefore Subway was reinstated and a sample of 21 restaurants’ data was collected.

### Data Collection

Menu data was collected directly from restaurants’ UK websites in February and March of 2024 (May 2024 for Subway). It was collected primarily through PDF menu documents or menu webpages. Where a restaurant provided multiple PDFs, data was only collected from those labelled as containing ‘core’ (or equivalent) menu items. Limited time offer menu items (e.g., seasonal) were only included in data collection if they were present in the ‘core’ menus. For pizza restaurants that offered multiple crust options, only nutrition information from the default option (e.g. classic crust) was collected, but all size variations (e.g. small, medium, large) were included.

Data in PDFs were extracted either by copying and pasting the data into Excel or by using an online data extraction software (Smallpdf (23)). Where data had to be extracted from menu webpages, we used the web scraping tool Octoparse (v8 desktop (24)) to extract the menu data into an Excel sheet. The extracted data was crosschecked against the source material by AOH. The data collection approach for each restaurant is detailed in S2 Table.

The data extracted included restaurant name, product name, nutritional information, and serving size, where available. Nutritional information was collected per 100g and per serving wherever given, and included kcal, kJ, fat, saturated fat, carbohydrates, sugar, protein, fibre, salt, and sodium.

### Categorising Menu Items

All menu items were categorised twice. First, author AOH assigned all menu items to one of the following 12 common subcategories: Pizzas, Burgers, Chicken, Other Mains, Children’s Meals, Salads, Sandwiches, Potato Sides, Other Sides, Breakfast Items, Desserts, and Sauces. This provided a consistent framework in which we could compare the nutritional content of similar menu items across restaurants. Hereafter, this categorisation will be referred to as an item’s ‘subcategory’. Categorisation criteria for each subcategory is detailed in S3 Table.

Menu items were also matched to the OOH food categories as defined by the technical guidance for the sugar, salt, and calorie reduction targets (8,10,11). The target-specific categories are not inclusive of all menu items and the guidance states that there should be no overlap between the calorie and sugar targets. Therefore, a menu item could be (i) ineligible for all three target types, (ii) eligible for only one of the three target types, (iii) eligible for a salt and calorie target but not a sugar target, or (iv) eligible for a salt and sugar target but not a calorie target. As the publication of the sugar targets preceded that of the calorie targets, menu items were categorised to a sugar target and not to a calorie target to avoid overlap.

A random 10% of menu items’ categorisations were checked by two co-authors (LB and HF) who had not been involved in the original categorisation process. A small subset of menu items was re-categorised and changes were applied to the complete dataset where relevant, followed by a final round of checking by author AOH.

We divided our 21 restaurants into the following five ‘restaurant types’, based on similarities in the focus of their menu portfolios: Burger, Chicken, Pizza, Sandwich, and Other Main. The 12 subcategories were also divided into ‘Mains’ and ‘Sides/Extras’, with ‘Desserts’ in its own category. Throughout our results we will report statistical comparisons between restaurants and subcategories within their groupings.

### Analysis

Data analyses were conducted in R Studio (version 4.3.2), using the following packages: readxl, dplyr, tidyr, stringr, tidyverse, zoo, writexl, reshape, ggplot2.

#### Missing Data

We excluded menu items where all three of the nutrients of interest (kcal, salt, and sugar), were missing. For menu items that were missing two or fewer of the nutrients of interest, where possible we calculated the missing values using the following formulas:

*Kcal = kJ / 4.184 (and vice versa)*

*Kcal = (Protein (g) * 4) + (Fat (g) * 9) + (Carbohydrate * 3.75)*

*Sodium (mg) = (Salt (g) * 1000) / 2.5 (and vice versa)*

In order to assess an item’s adherence to the Government’s reduction targets, nutrition information was needed per 100g for the sugar targets, per serving for the calorie targets, and in both formats for the salt targets. If per 100g, per serving, or serving size information was missing, the missing value was calculated from the provided information using the following formulas:

*Per 100g = Per serving / (Serving Size / 100)*

*Per Serving = (Serving size / 100) * Per 100g*

*Serving Size = (100 / Per 100g) * Per Serving*

Here, ‘Serving Size’ refers to the weight of the menu item in grams, and ‘Per 100g’ and ‘Per Serving’ refer to nutrient content (e.g., salt content in 100g or in a single serving of a menu item).

Where it was not possible to calculate serving size (only per 100g or only per serving nutrient information was provided), the mean serving size for the menu item’s subcategory was used instead. For example, if the serving size for a burger was not provided, the mean serving size of menu items within the ‘Burger’ subcategory was used instead.

#### Average Nutrient Content

The mean and median kcal, sugar, salt, and fat content was calculated for each restaurant and subcategory. Averages were calculated using nutrient information provided per 100g, per reported serving size, and per subcategory average serving size. ‘Reported serving size’ refers to the serving size of a menu item as reported by the restaurant, whereas ‘subcategory average serving size’ refers to the average serving size of menu items within a subcategory (e.g., the average serving size for menu items within the ‘Burger’ subcategory).

For Pizzas, average nutrient content per serving was calculated using the serving sizes provided by individual restaurants. Papa John’s provided per serving information per slice, Pizza Hut and Domino’s provided per serving information per person sharing (e.g., medium pizza is shared between two people, so one pizza would count as two servings), and the remaining restaurants provided per serving information per whole pizza.

Kruskal-Wallis tests with the independent variable of restaurant or subcategory and the dependent variables of kcal, salt, and sugar per 100g (separate tests for each nutrient), were conducted to determine whether nutrient content per 100g significantly differed between restaurants and subcategories. Post-hoc Dunn tests were conducted to compare restaurants and subcategories within groupings (e.g., compare restaurants within the ‘Pizza’ restaurant group, or compare subcategories within the ‘Mains’ group), with Bonferroni correction applied to account for the number of comparisons (e.g., four restaurants within the ‘Pizza’ restaurant group, meaning six comparisons were conducted, and therefore *p*=0.050/6 = 0.008).

#### Adherence to Targets

The sugar, salt, and calorie content of each product was compared to their matched category’s target value. S4 Table shows the range of target values that menu items had to meet, within each subcategory. Where a menu item’s sugar, salt, or calorie content was equal to or lower than the target value, they were deemed to have met the target. The proportion of each restaurant and subcategory’s menu items that met the targets was expressed as a percentage of their total number of menu items eligible for the respective target.

Logistic regressions with the predictor variable of restaurant and the binary outcome of ‘target met’ (0 for ‘no’ and 1 for ‘yes’) were used to determine whether the restaurant that offered a menu item, could significantly predict whether that menu item would meet its target. Separate regressions were conducted for each of the three target types (sugar, salt, and calories), and for a fourth combined measure of ‘all applicable’ targets. A menu item was said to meet ‘all applicable’ targets if they met all of the targets that they were eligible for (e.g., if a menu item had a calorie and a salt target, they would need to meet both in order to meet ‘all applicable’ targets). Separate regressions were conducted for each restaurant grouping, such that restaurants with similar menu portfolios were entered into the same model. Bonferroni corrections were applied to each model to control for multiple comparisons, where each restaurant was compared to the reference restaurant (e.g., four pizza restaurants, meaning three comparisons to the reference group were conducted, and therefore *p*=0.05/3=0.017).

#### Sensitivity Analysis

We repeated the primary analyses for mean nutrient content and target adherence but with limited-time offer menu items that featured on the main menu, excluded.

#### Deviations from Protocol

For our analysis of average nutrient content, we had planned to use factorial ANOVAs with the independent variables of restaurant and subcategory. Firstly, we used Kruskal-Wallis tests instead of ANOVAs as the data was not normally distributed. Secondly, due to the variation across restaurants in the number of menu items belonging to each subcategory, instead of including subcategory as an additional independent variable, we grouped the restaurants based on their prominent ‘main meal’ subcategory and reported pairwise comparisons between restaurants within the same group.

For our second sensitivity analysis, we originally planned to repeat the primary analysis for target adherence but using ‘as sold’ nutrition information instead of ‘per serving’ information. For example, if a restaurant reports that a menu item contains two servings, our primary analysis would have assessed target adherence based on a single serving (being ‘per serving’), whilst our sensitivity analysis would assess it as the whole menu item (‘as sold’). The aim of this analysis was to remove the influence of individual restaurants’ reporting of serving size, to allow for a consistent assessment of nutritional quality across restaurants. On examination of the data, we found for Pizzas in particular, the suggested serving sizes and reporting of per serving nutrition information lacked consistency across restaurants. For example, Papa John’s provided their nutrition information per slice without explicitly stating how many slices equate to a single serving, whilst Pizza Express provided information per whole pizza with no suggestions for how many servings it contained. The technical guidance for the salt and calorie targets attempts to account for this, with the salt targets being applied per slice or per whole pizza depending on the style of pizza (takeaway or Italian-style), and the calorie targets being applied per serving for ‘sharing’ pizzas (defined as large and above, or 11.5” and above). Our analysis followed this guidance, with assumptions for serving size being made to apply the calorie targets where information was not provided. For example, Papa John’s large and extra-large pizzas were assumed to contain three and four servings respectively, as they did not provide their own suggestions, and Italian-style pizzas were assumed to be a single serving. For analyses with the outcome of average nutrient content per serving, serving sizes for Pizzas were as provided by the restaurants.

We planned to conduct a sensitivity analysis in which we would repeat all primary analyses, but for menu items that did not have a serving size reported, we would use an applicable serving size from the Food Standard’s Agency ‘Food Portion Sizes’ handbook (25). Upon further investigation, we concluded that the suggested serving sizes from this handbook would not be applicable to out-of-home menu items, as they were often provided per meal component rather than complete meal (e.g., suggested serving size provided for a burger patty and a bun, rather than a whole burger), and therefore this analysis was not conducted.

The planned exploratory analysis comparing the same menu items reported in 2022 with 2024 was not conducted due to the small sample size of products present on menus in both years.

As the targets are food only, and the same top-selling brands of drinks appear on the majority of restaurant’s menus, drinks were excluded.

The planned NPM analysis will be presented in a separate paper.

## Results

A total of 3099 menu items across 21 restaurants were included in this study. Originally, 5435 menu items were collected. Of the 2336 items excluded, 453 were duplicates, 103 did not have the required nutritional information, 825 were pizzas with non-default crust options, and 955 were drinks. One menu item was missing a sugar value, so this item was excluded from analyses where sugar content is the outcome.

The mean number of menu items per restaurant was 148, although this varied widely from 40 items for Burger King to 330 items for Pizza Hut (Table 1). The number of subcategories offered by each restaurant ranged from 5 for Starbucks and Burger King, to 11 for Leon.

**Table 1.**
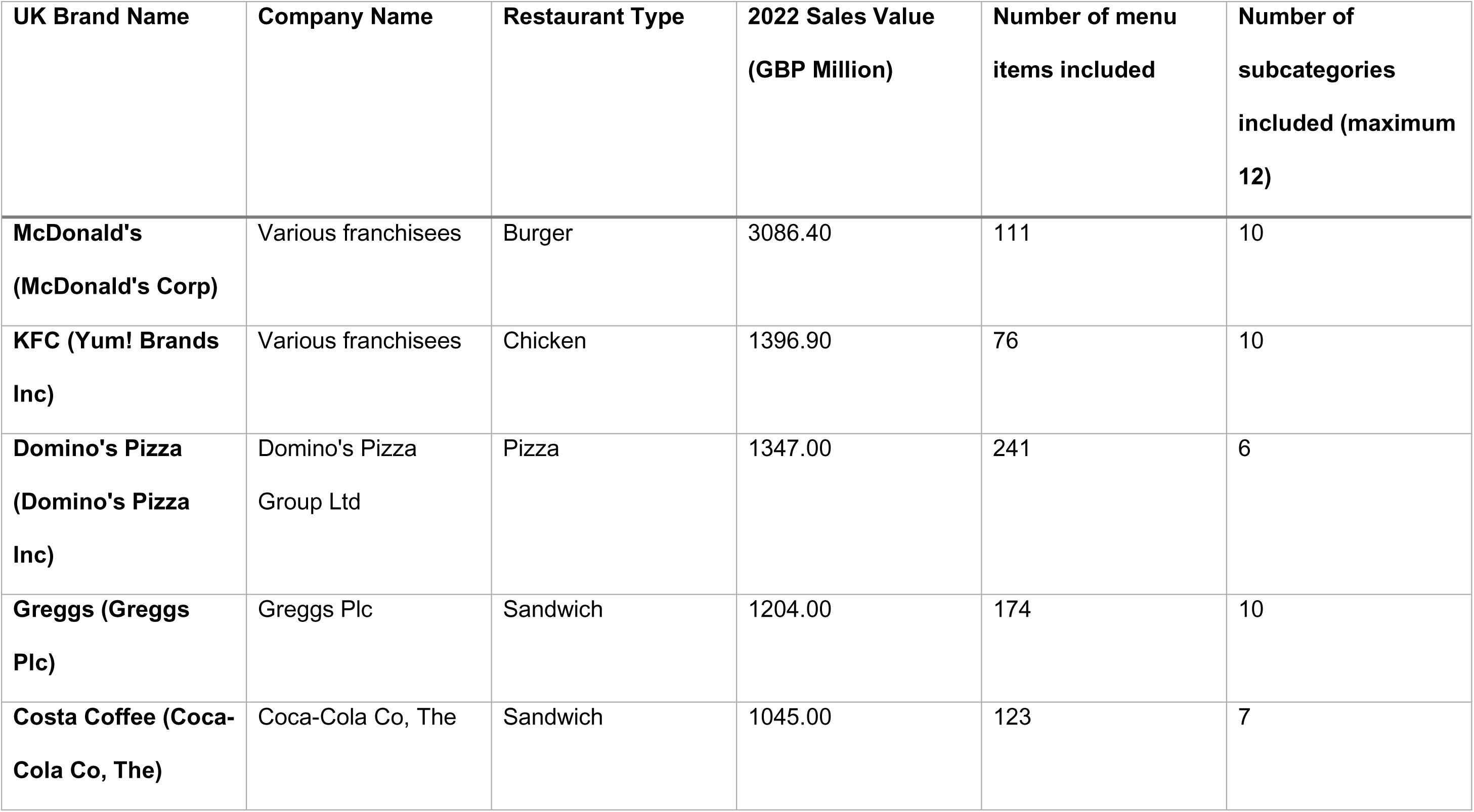
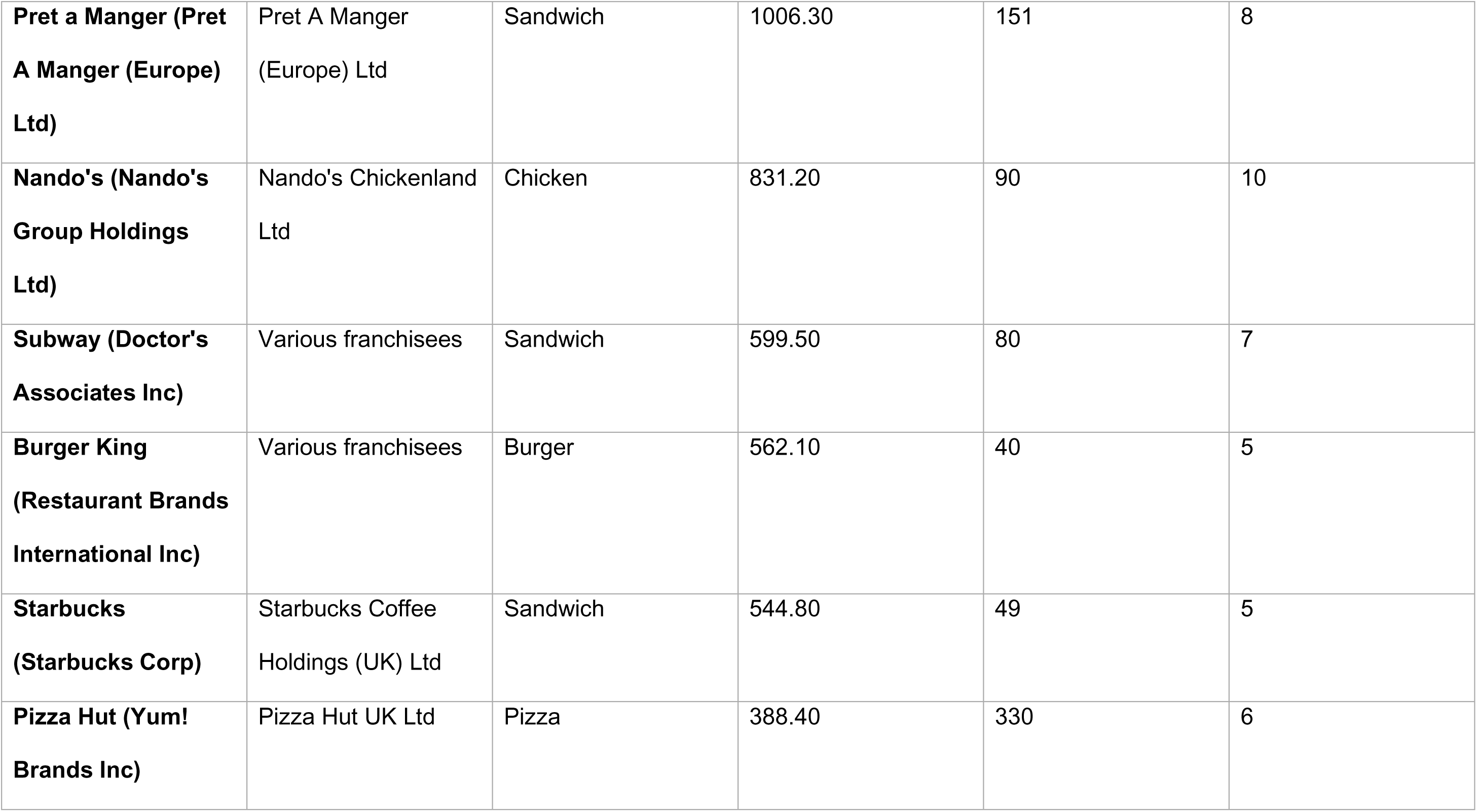
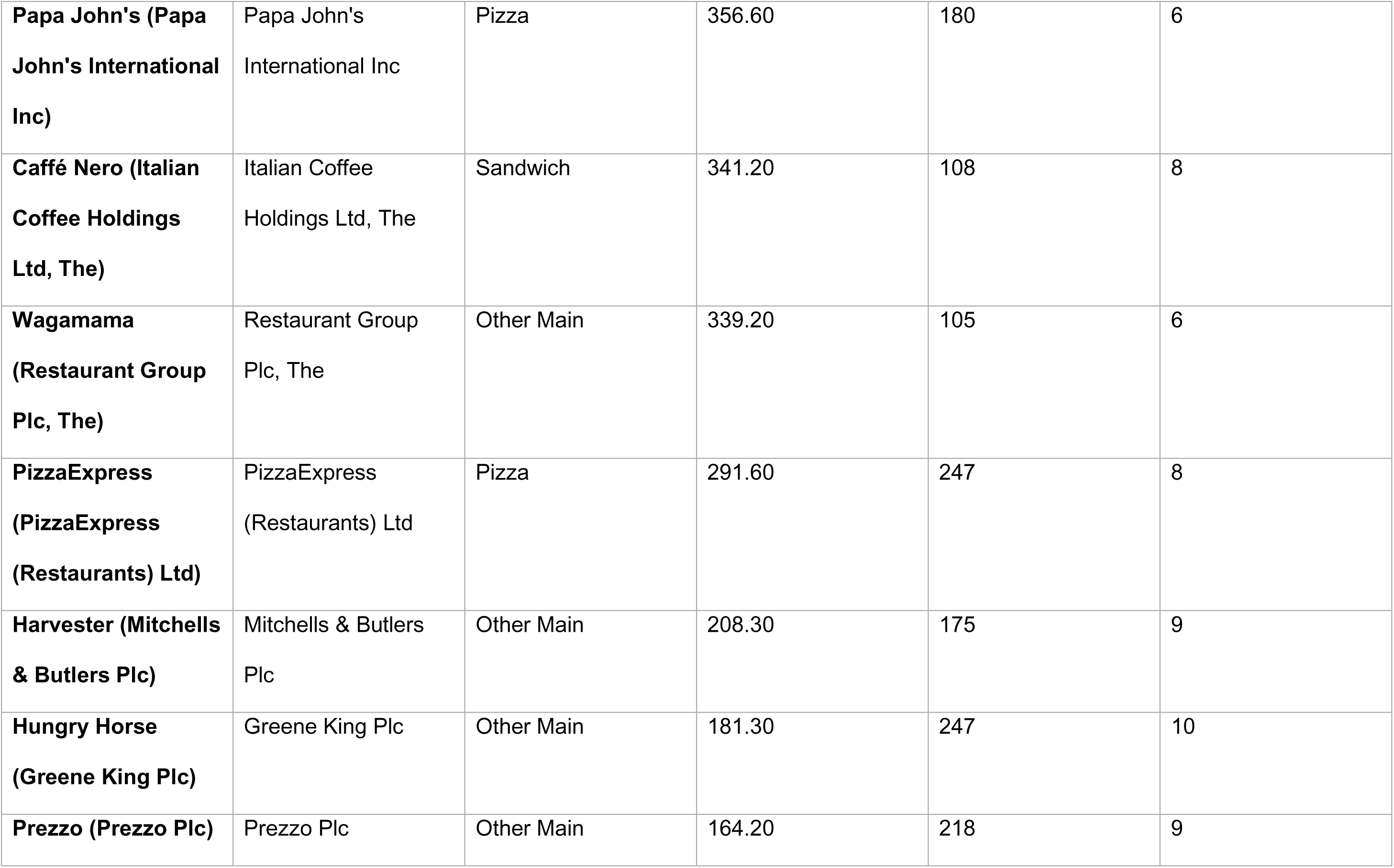
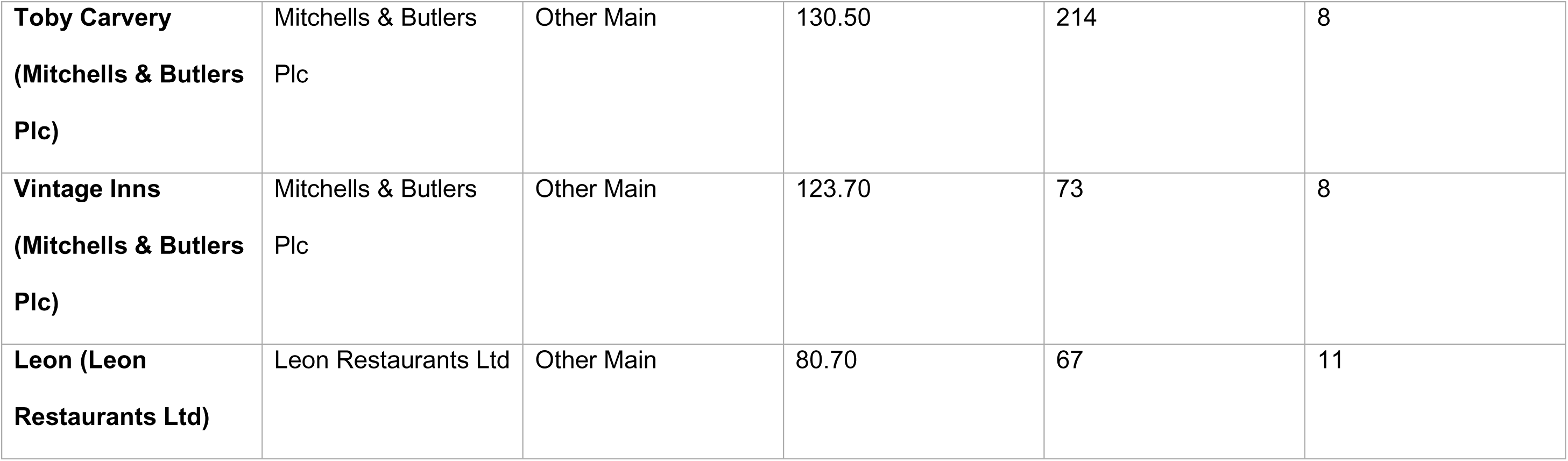
A summary of characteristics for each restaurant, including their affiliated company, type, 2022 sales value, number of menu items included, and number of subcategories included. Restaurants presented in descending order by 2022 Sales Value.

| **UK Brand Name** | **Company Name** | **Restaurant Type** | **2022 Sales Value**<br>**(GBP Million)** | **Number of menu**<br>**items included** | **Number of**<br>**subcategories**<br>**included (maximum**<br>**12)** |
| --- | --- | --- | --- | --- | --- |
| **McDonald's**<br>**(McDonald's Corp)** | Various franchisees | Burger | 3086.40 | 111 | 10 |
| **KFC (Yum! Brands**<br>**Inc)** | Various franchisees | Chicken | 1396.90 | 76 | 10 |
| **Domino's Pizza**<br>**(Domino's Pizza**<br>**Inc)** | Domino's Pizza<br>Group Ltd | Pizza | 1347.00 | 241 | 6 |
| **Greggs (Greggs**<br>**Plc)** | Greggs Plc | Sandwich | 1204.00 | 174 | 10 |
| **Costa Coffee (Coca-**<br>**Cola Co, The)** | Coca-Cola Co, The | Sandwich | 1045.00 | 123 | 7 |
| **Pret a Manger (Pret A Manger (Europe) Ltd)** | Pret A Manger (Europe) Ltd | Sandwich | 1006.30 | 151 | 8 |
| **Nando's (Nando's Group Holdings Ltd)** | Nando's Chickenland Ltd | Chicken | 831.20 | 90 | 10 |
| **Subway (Doctor's Associates Inc)** | Various franchisees | Sandwich | 599.50 | 80 | 7 |
| **Burger King (Restaurant Brands International Inc)** | Various franchisees | Burger | 562.10 | 40 | 5 |
| **Starbucks (Starbucks Corp)** | Starbucks Coffee Holdings (UK) Ltd | Sandwich | 544.80 | 49 | 5 |
| **Pizza Hut (Yum! Brands Inc)** | Pizza Hut UK Ltd | Pizza | 388.40 | 330 | 6 |
| **Papa John's (Papa John's International Inc)** | Papa John's International Inc | Pizza | 356.60 | 180 | 6 |
| **Caffé Nero (Italian Coffee Holdings Ltd, The)** | Italian Coffee Holdings Ltd, The | Sandwich | 341.20 | 108 | 8 |
| **Wagamama (Restaurant Group Plc, The)** | Restaurant Group Plc, The | Other Main | 339.20 | 105 | 6 |
| **PizzaExpress (PizzaExpress (Restaurants) Ltd)** | PizzaExpress (Restaurants) Ltd | Pizza | 291.60 | 247 | 8 |
| **Harvester (Mitchells & Butlers Plc)** | Mitchells & Butlers Plc | Other Main | 208.30 | 175 | 9 |
| **Hungry Horse (Greene King Plc)** | Greene King Plc | Other Main | 181.30 | 247 | 10 |
| **Prezzo (Prezzo Plc)** | Prezzo Plc | Other Main | 164.20 | 218 | 9 |
| **Toby Carvery**<br>**(Mitchells & Butlers**<br>**Plc)** | Mitchells & Butlers<br>Plc | Other Main | 130.50 | 214 | 8 |
| **Vintage Inns**<br>**(Mitchells & Butlers**<br>**Plc)** | Mitchells & Butlers<br>Plc | Other Main | 123.70 | 73 | 8 |
| **Leon (Leon**<br>**Restaurants Ltd)** | Leon Restaurants Ltd | Other Main | 80.70 | 67 | 11 |

Some restaurants had large menus that focused on a smaller number of subcategories, such as Pizza Hut and Domino’s, whilst others had smaller menus with more diverse types of items, such as Leon (Figure 1).

**Figure 1.**
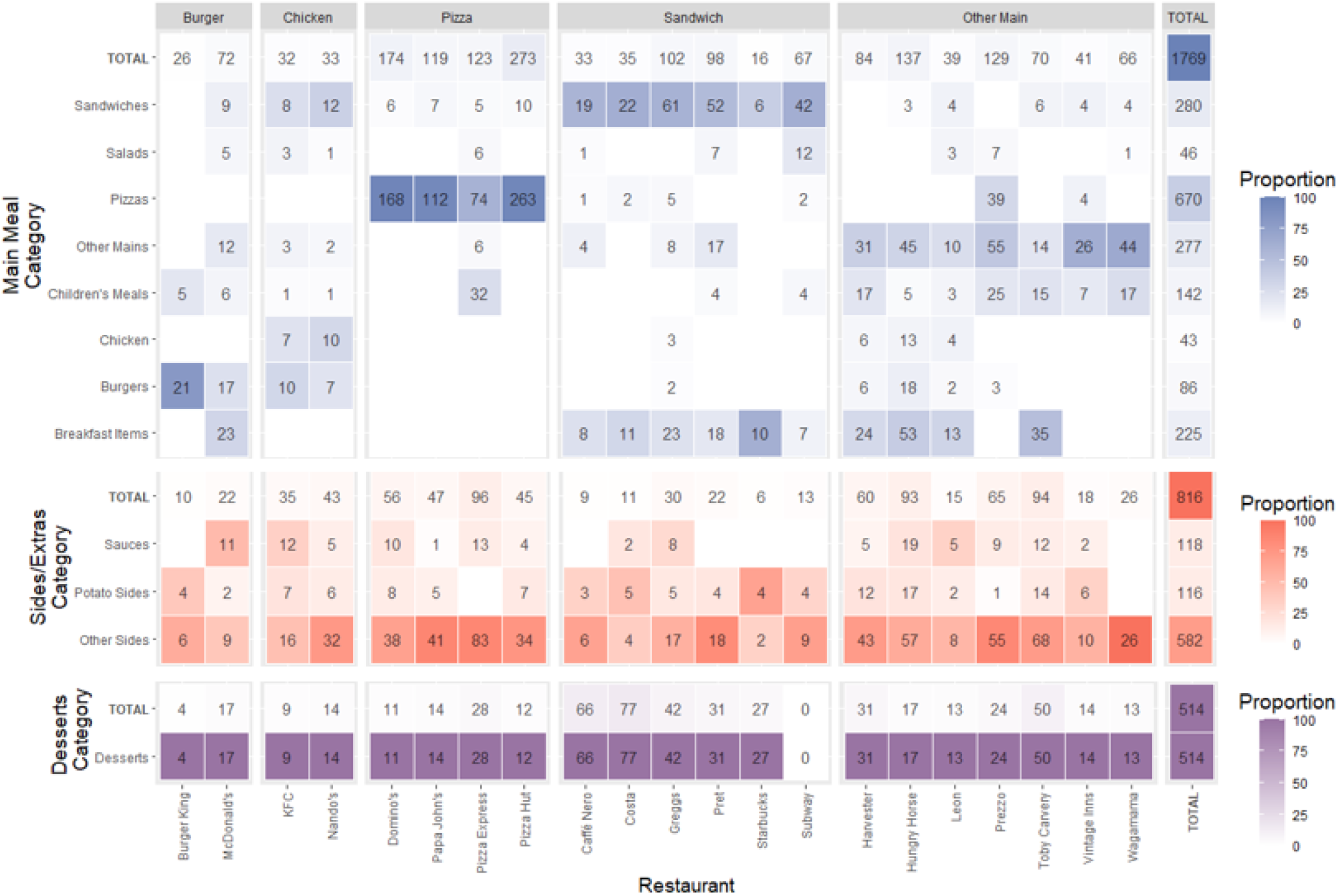
The number and proportion of menu items by restaurant and subcategory. Notes: KFC and Nando’s were characterised as ‘Chicken’ restaurant type despite majority of main meal menu items belonging to ‘Sandwich’ or ‘Burger’ subcategories, as these menu items were majority chicken burgers and chicken wraps. The same principle was applied to restaurants with the majority of their main menu items being ‘Breakfast Items’.

### Average Nutrient Content

#### By Subcategory

Per 100g, mean nutrient content across all menu items was 280kcal, 1.1g salt, and 9.5g sugar. Per serving, mean nutrient content across all menu items was 450kcal, 2.0g salt, and 10.9g sugar.

Desserts had the highest mean calorie (410kcal) and sugar (34.3g) content per 100g across all subcategories, whilst Sauces had the highest salt content per 100g (2.2g) (Figure 2). Per serving, Other Mains had the highest calorie content (756kcal) and joint highest salt content with Pizzas (3.4g), and Desserts had the highest sugar content (26.2g) (Figure 3). Means, Medians, and Standard Deviations for nutrient content per 100g, per reported serving, and per subcategory average serving, for each subcategory, are found in S5-8 Tables.

**Figure 2.**
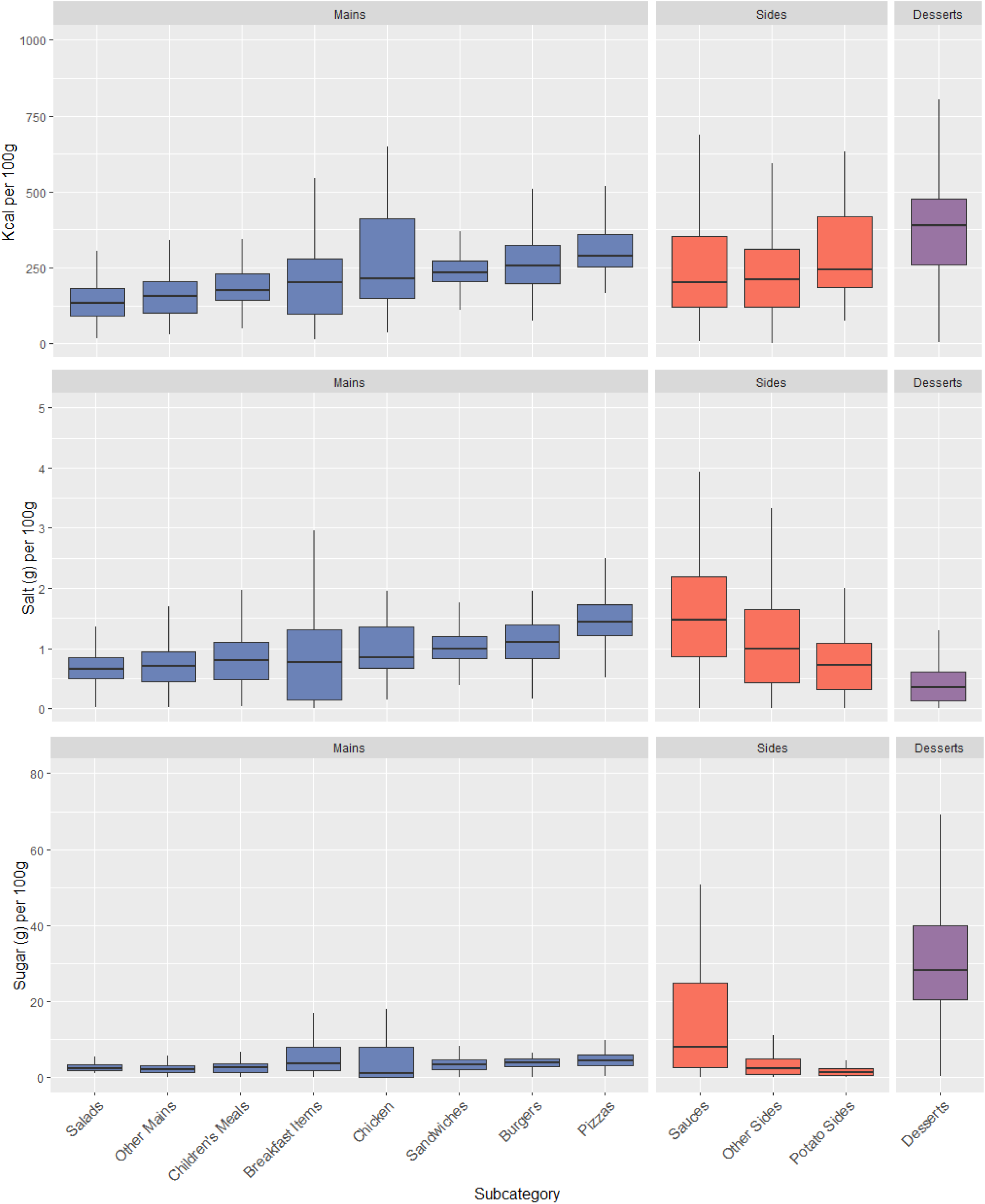
The distribution of kcal, salt, and sugar content <u>per 100g</u> for menu items by subcategory. Notes: Subcategories are ordered ascendingly by median Kcal per 100g. Mid-lines represent the median, the box represents the inter-quartile range, and whiskers represent the range.

**Figure 3.**
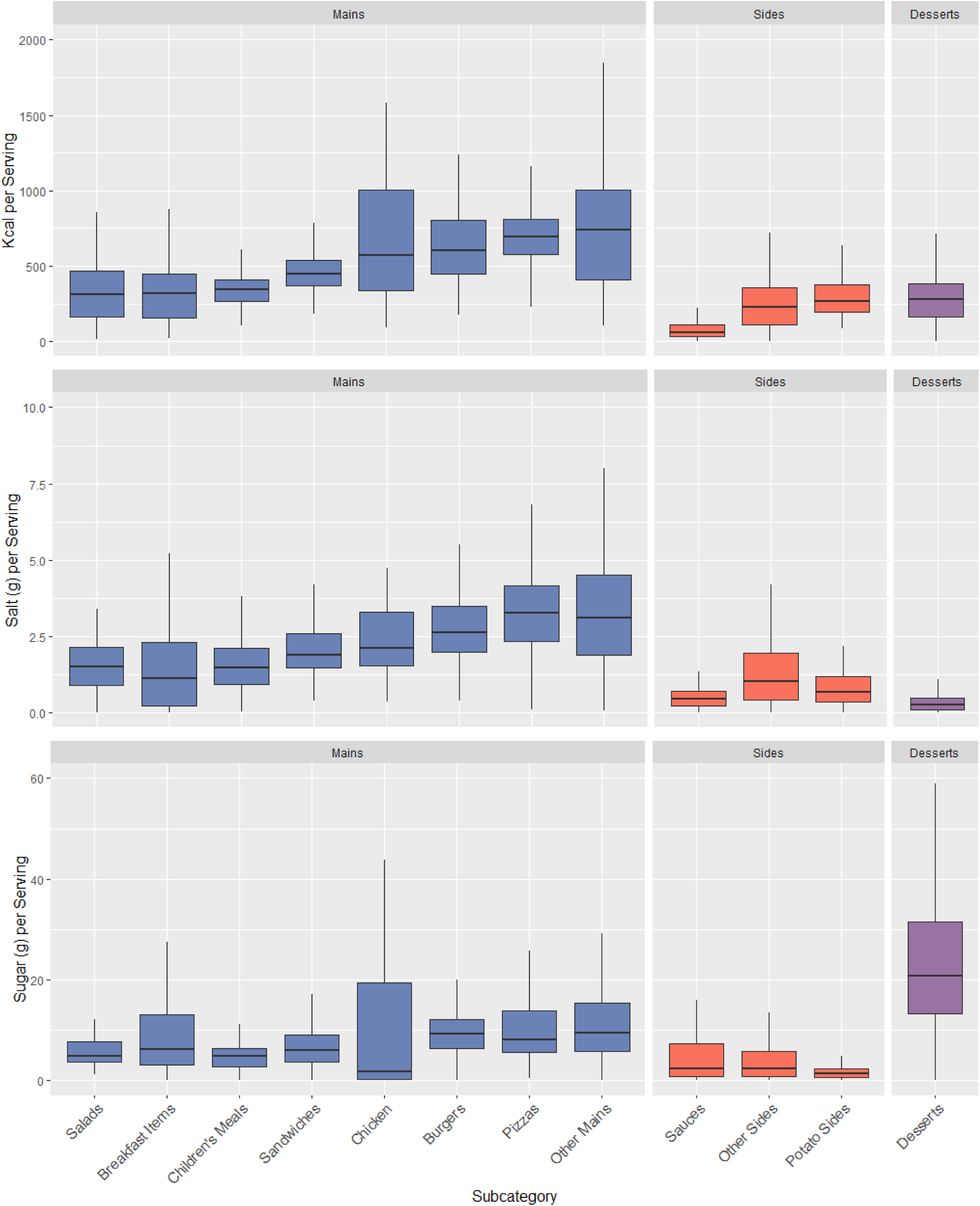
The distribution of kcal, salt, and sugar content <u>per serving</u> for menu items by subcategory. Notes: Subcategories are ordered ascendingly by median Kcal per serving. Mid-lines represent the median, the box represents the inter-quartile range, and whiskers represent the range.

#### Difference testing

Kruskal-Wallis tests found sugar, salt, and kcal per 100g to vary significantly by subcategory (df=20, *p*<.001, sugar X^2^= 1427.50, salt X^2^= 1054.50, kcal X^2^=746.31). Comparing between the main meal subcategories, Pizzas had the highest median sugar, salt, and calorie content per 100g, with Dunn tests finding significant differences between Pizzas and all remaining categories for salt (*p*<.001) per 100g, all categories except Burgers for kcal (*p<.*001), and all categories except for Breakfast Items (*p*=.006) and Burgers (*p*=.059) for sugar per 100g. Salads had the lowest median calorie and salt content per 100g, differing significantly to all remaining mains categories except for Children’s Meals (*p*=.037) and Other Mains (*p*=.395) for kcal per 100g, but only differing significantly in salt per 100g to Burgers, Pizzas, and Sandwiches (*p*<.001). Chicken had the lowest sugar content per 100g, but only differed significantly to Breakfast Items and Pizzas (*p*<.001). Comparing between the sides/extras subcategories, Sauces had the highest median salt and sugar content per 100g, differing significantly to both Potato Sides and Other Sides (*p*<.001), but had the lowest median calorie content per 100g, differing significantly from Potato Sides only (*p*=.014). Potato Sides had the highest calorie content per 100g, differing significantly to both Sauces (*p*=.014) and Other Sides (*p*<.001), but had the lowest salt and sugar content per 100g, differing significantly again to both Sauces and Other Sides (*p*<.001). Pairwise comparisons between subcategories in median sugar, salt, and calorie content per 100g, are reported in S9 Table.

### By Restaurant

Caffé Nero had the highest mean calorie content per 100g (368kcal) across all restaurants, whilst Prezzo had the highest salt content (1.9g), and Costa had the highest sugar content (22.0g) (Figure 4). Per serving, Prezzo had the highest calorie (644kcal) and salt content (3.7g) and Harvester had the highest sugar content (17.2g) (Figure 5). Means, Medians, and Standard Deviations for nutrient content per 100g, per reported serving, and per subcategory average serving, for each restaurant, are found in S10-13 Tables.

**Figure 4.**
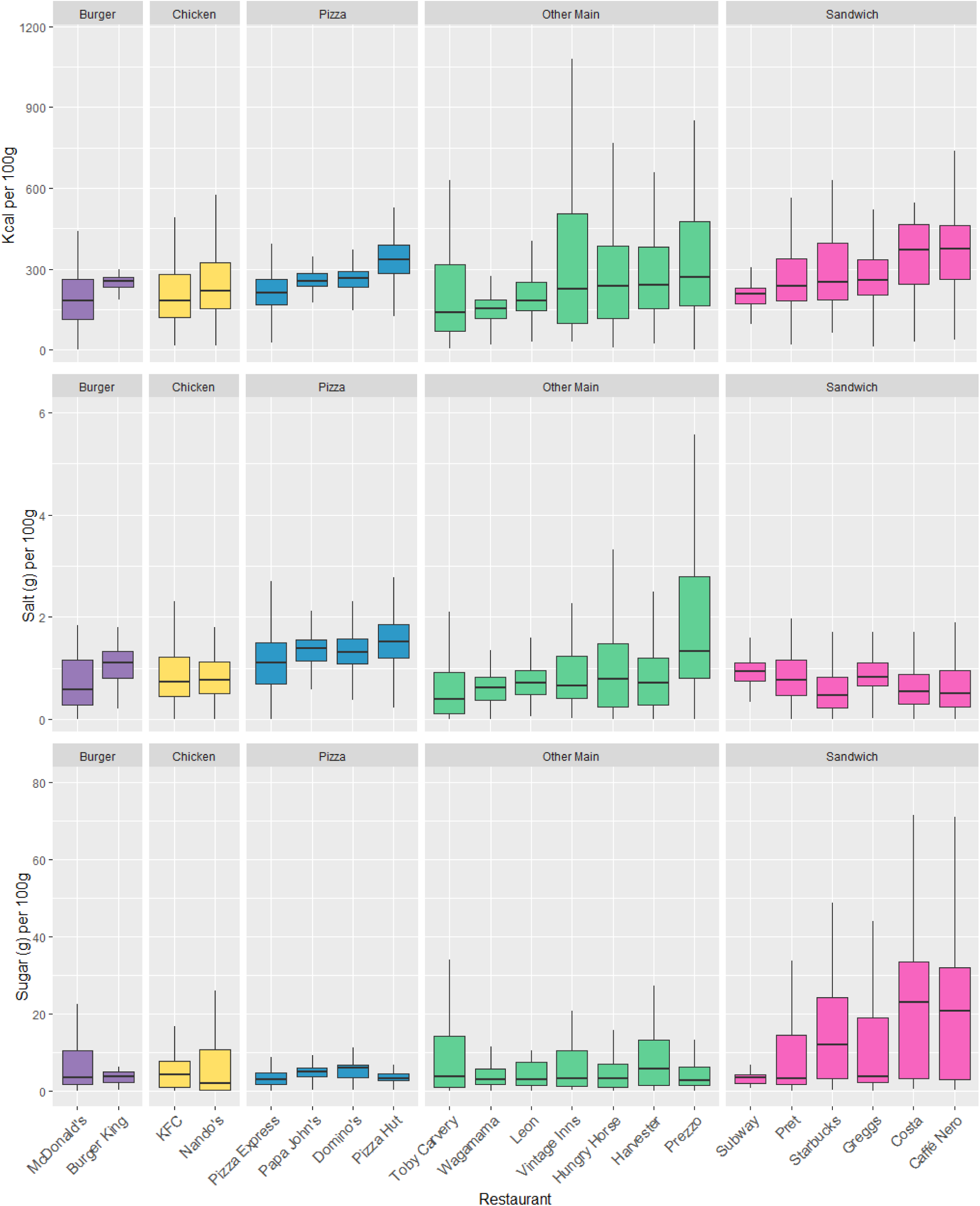
The distribution of kcal, salt, and sugar content per 100g for menu items by restaurant. Notes: Restaurants are ordered ascendingly by median Kcal per 100g. Mid-lines represent the median, the box represents the inter-quartile range, and whiskers represent the range.

**Figure 5.**
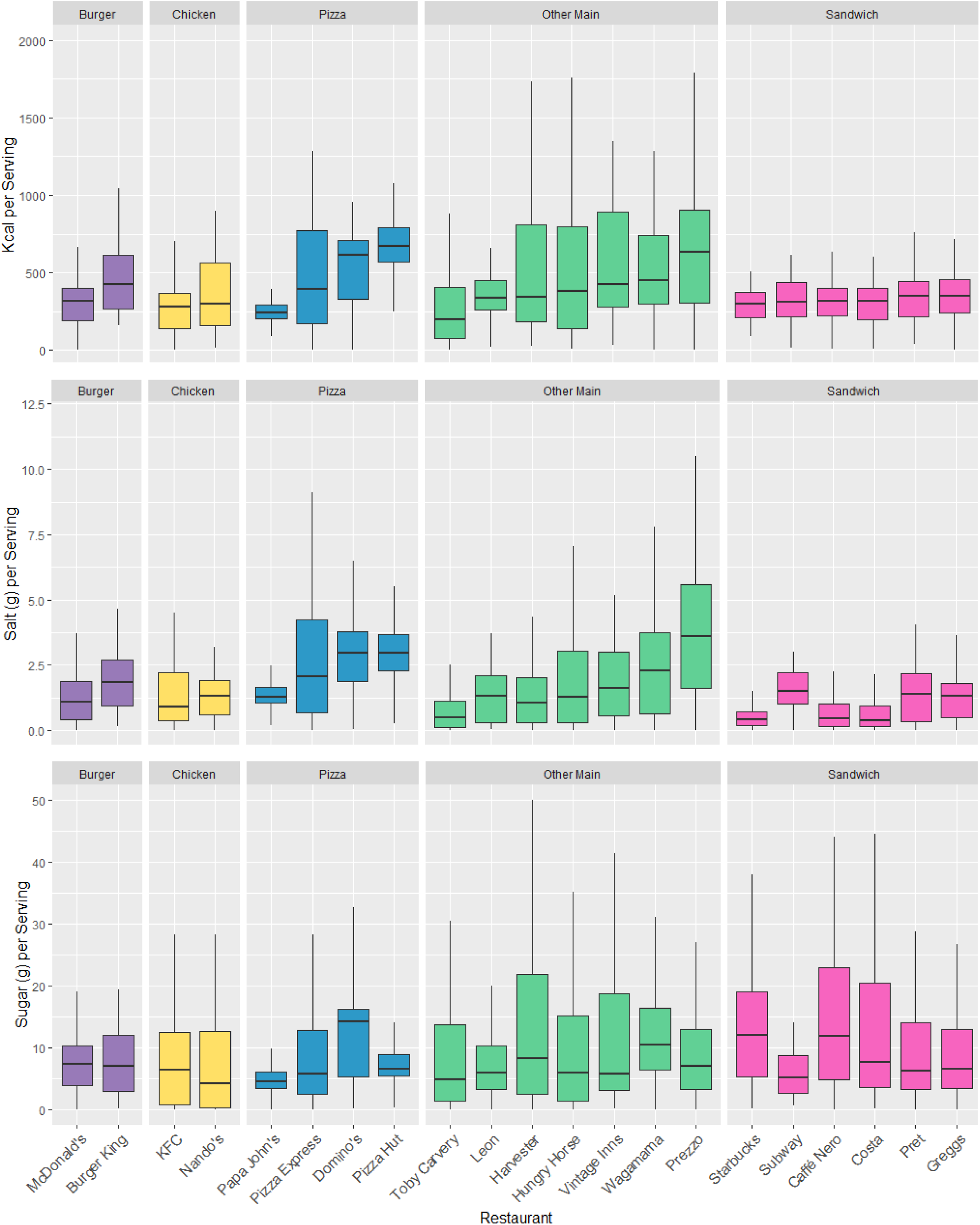
The distribution of kcal, salt, and sugar content per serving for menu items by restaurant. Notes: Restaurants are ordered ascendingly by median Kcal per serving. Mid-lines represent the median, the box represents the inter-quartile range, and whiskers represent the range.

Averaging across menu items within restaurant groups, the Sandwich group had the highest mean calorie (293kcal) and sugar content per 100g (13.4g), and the Pizza group had the highest mean salt content (1.4g). The Pizza restaurant group had the highest mean calorie (515 kcal) and salt content per serving (2.6g), and the Other Mains group had the highest mean sugar content (12.6g). Means, Medians, and Standard Deviations for nutrient content per 100g, per reported serving, and per subcategory average serving, for each restaurant group, are found in S14-17 Tables.

#### Difference testing

Kruskal-Wallis tests found sugar, salt, and kcal per 100g to vary significantly by restaurant (df=20, *p*<.001, sugar X^2^=229.03, salt X^2^=711.22, kcal X^2^=497.76). Comparing between the Pizza restaurants, Pizza Hut had the highest median calorie content per 100g, differing significantly to the each of the three other Pizza restaurants (*p*<.001), and the highest median salt content per 100g, but differing significantly to only Pizza Express (*p*<.001). Domino’s had the highest median sugar content per 100g, but only differed significantly to Pizza Express and Pizza Hut (*p*<.001). Nando’s had higher calorie and salt content per 100g than KFC, whilst KFC had higher median sugar content per 100g, however none of these differences were significant (*p*=.061-.899). Burger King had higher median sugar, salt, and calorie content per 100g than McDonald’s, but this difference was only significant for calories (*p*=.003) and salt (*p*=.006). Comparing between the Sandwich restaurants, Caffé Nero had the highest median calorie content per 100g, differing significantly to all other restaurants except Costa (*p*=.605). Subway had the highest median salt content per 100g, differing significantly to all other restaurants except Greggs (*p*=.551) and Pret (*p*=.076). Costa had the highest median sugar content per 100g, but only differed significantly to Pret, Greggs, and Subway (*p*<.001). Comparing between the Other Mains restaurants, Prezzo had the highest median salt content per 100g, differing significantly to all other restaurants (*p*<.001), and calorie content per 100g, but differing significantly to Leon, Wagamama, and Toby Carvery only (*p*<.001). Finally, Harvester had the highest median sugar content per 100g, but differed significantly to Hungry Horse and Prezzo only (*p*<.001). Pairwise comparisons between restaurants in median sugar, salt, and calorie content per 100g, are reported in S18 Table.

### Target Adherence

Across all restaurants and subcategories, 61% of menu items met their calorie targets, 58% met their salt targets, 36% met their sugar targets, and 43% met all of their applicable targets.

#### By Subcategory

Six out of the 12 subcategories had over 50% of their menu items meeting all applicable targets. Salads had the highest adherence to all applicable targets at 96% but were only eligible for the calorie targets (therefore having 96% adherence for calorie targets as well). Breakfast Items had the second highest adherence to all applicable targets at 66%, whilst being eligible for all three target types. Excluding Other Sides and Children’s Meals (which had 100% adherence to the sugar targets but each had only one eligible item), the highest sugar target adherence was seen for Breakfast Items at 74%, which also had the highest salt target adherence at 82% (Figure 6).

**Figure 6.**
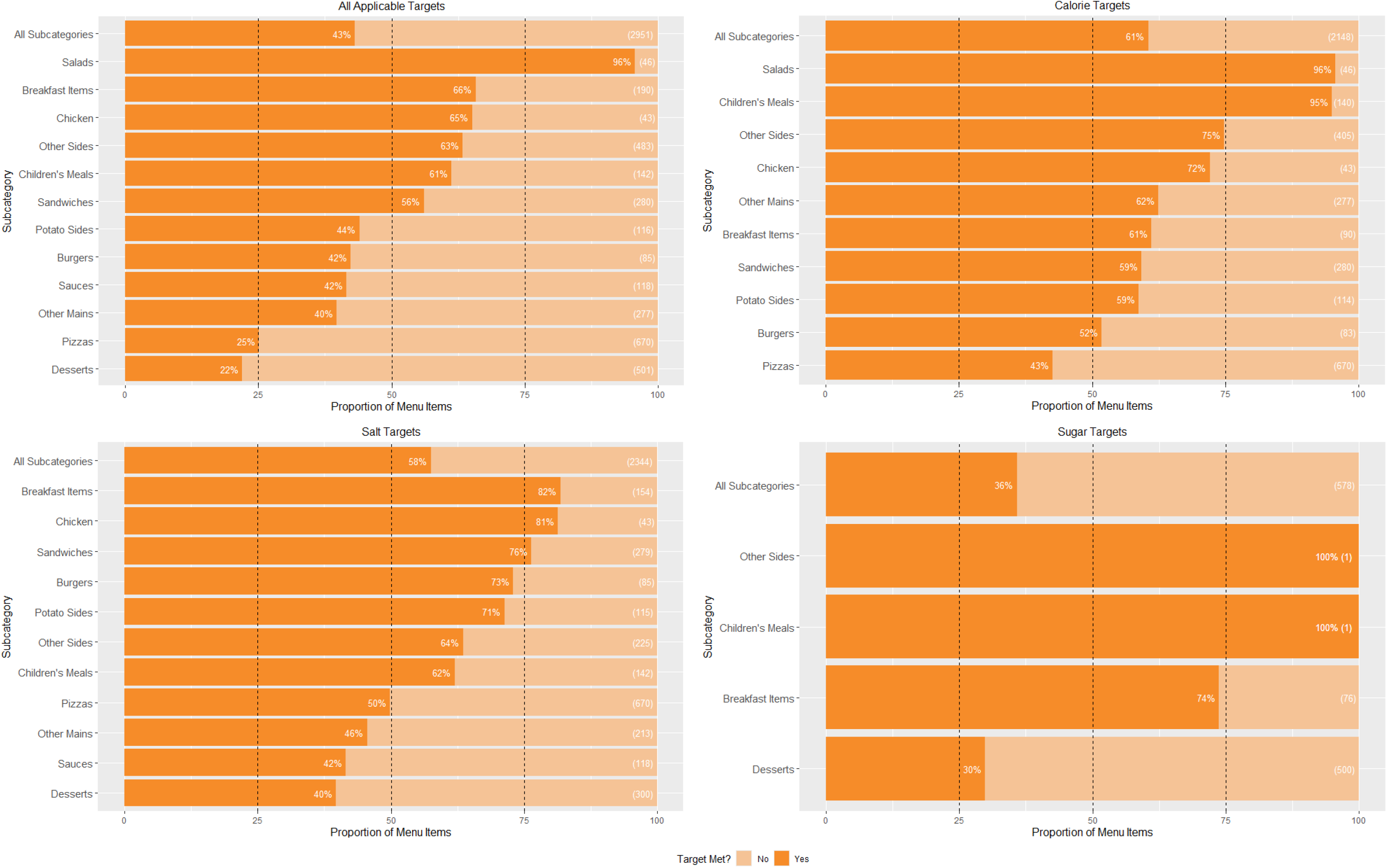
The proportion of menu items that met all applicable, calorie, salt, and sugar targets, by subcategory. Notes: Values in brackets show the total number of eligible menu items in that subcategory. Subcategories in descending order by proportion of menu items meeting the respective target.

### By Restaurant

Nine of the 21 restaurants had over 50% of their menu items meeting all applicable targets (Figure 7).

**Figure 7.**
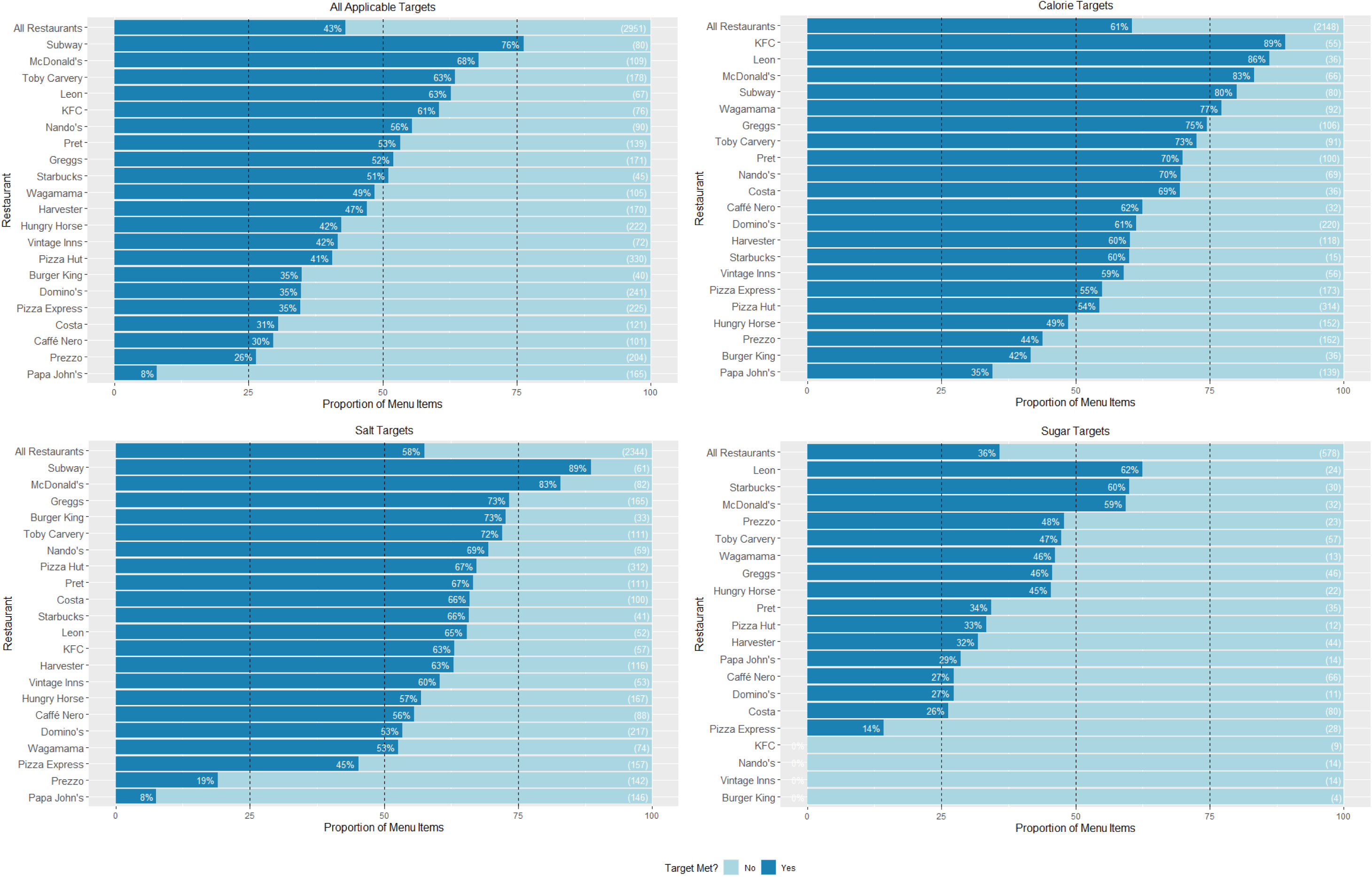
The proportion of menu items that met all applicable, calorie, salt, and sugar targets, by restaurant. Notes: Values in brackets show the total number of eligible menu items for that restaurant. Restaurants in descending order by proportion of menu items meeting the respective target.

The Pizza restaurant group had the lowest combined adherence to all applicable targets at 32%, to the salt targets at 49%, and to the calorie targets at 53%. Adherence to sugar targets was based on lower product numbers, with the Chicken restaurant group having the lowest adherence at 0%, but with only 23 eligible items. The Pizza restaurant group was the second lowest adhering to the sugar targets at 23%, with 65 eligible items.

The Burger restaurant group had the highest combined adherence to all applicable targets at 59%, and to the salt targets at 80%, whilst the Chicken group had the highest combined adherence to the calorie targets at 78%. The Burger restaurant group had the highest adherence to the sugar targets at 53%, but with only 36 eligible items, of which 32 were from McDonald’s. Full results for adherence to each target type by restaurant group can be found in S19 Table.

Logistic regressions revealed that a menu item from McDonald’s had a significantly higher likelihood of meeting their calorie and all applicable targets than a menu item from Burger King (calorie: *p*<.001, OR=7.00; all applicable: *p*<.001, OR=3.93). A menu item from KFC had a significantly higher likelihood of meeting their calorie target than a menu item from Nando’s (*p*=.012, OR=3.57). A menu item from any of the Other Mains restaurants had a significantly higher likelihood of meeting their salt target and all applicable targets than a menu item from Prezzo (salt: all *p*<.001, OR range=4.75-10.99; all applicable: Vintage Inns *p*=.017, other restaurants all *p*<.001, OR range=1.98-4.83), but only menu items from Harvester, Toby Carvery, Wagamama and Leon, had a significantly higher likelihood of meeting their calorie target than a menu item from Prezzo (Harvester *p*=.007, other restaurants all *p*<.001, OR range=1.94-7.95). A menu item from any of the three remaining Pizza restaurants had a significantly higher likelihood of meeting their calorie, salt, and all applicable targets than a menu item from Papa John’s (all *p*<.001, calorie OR range=2.27- 3.01, salt OR range=10.12-25.27, all applicable OR range=6.20-7.99). A menu item from Greggs or Subway had a significantly higher likelihood of meeting their salt target than a menu item from Caffé Nero (Greggs: *p*=.005, OR=2.19; Subway: *p*<.001, OR=6.14). A menu item from Starbucks had a significantly higher likelihood of meeting their sugar target than a menu item from Costa (*p*=.001, OR=4.21). Finally, a menu item from Greggs, Pret, or Subway had a significantly higher likelihood of meeting all applicable targets than a menu item from Caffé Nero (all *p*<.001, OR=2.57-7.60). Outcomes from the logistic regression models are reported in S20 Table.

### Sensitivity Analysis – Excluding Limited Time Offer Menu Items

We excluded 37 limited time offer menu items, spanning four restaurants (McDonald’s, Burger King, Pret, and KFC) and eight unique subcategories, for this analysis. Dunn tests comparing median calorie, salt, and sugar content per 100g across restaurants largely mirrored the primary analysis, except for KFC having significantly higher sugar content per 100g than Nando’s with limited time menu items excluded (*p=*.012). Pairwise comparisons for this sensitivity analysis are reported in S21 Table.

Logistic regressions did not find menu items from KFC to have a significantly higher likelihood of meeting their calorie target than menu items from Nando’s (*p*=.078), but all other findings mirrored the primary analysis. Outcomes from the logistic regressions for this sensitivity analysis are reported in S22 Table.

## Discussion

This study shows that only 43% of menu items from the highest-grossing UK restaurant chains met all of their reduction targets, suggesting there has been little engagement from the OOH sector with the reduction programmes. The majority of menu items met the calorie (61%) and salt (58%) reduction targets, however only 36% of menu items met sugar targets. Heterogeneity in adherence was observed across food categories, with Desserts having the lowest proportion of menu items meeting all applicable targets at 22%, and Salads the highest at 96%. Heterogeneity in target adherence was also observed across restaurants, with Papa John’s having the lowest proportion of menu items that met all their applicable targets at 8%, and Subway the highest at 76%. A similar number of restaurants met over 50% of their calorie and salt targets (17/21 and 18/21, respectively), but salt target adherence varied much more widely from 8-89%, compared to 35-89% for calories.

Subcategories were not always consistent in their performance across the target types, for example, Children’s Meals had 95% of menu items meeting calorie targets, but 62% meeting salt targets.

### Strengths and Limitations

This study is the first to evaluate all three reduction programmes in a single study, allowing us to compare engagement across the targets, both by individual company and overall. By providing an overview of restaurants’ whole menus, we were able to demonstrate that restaurants with similar menu portfolios displayed variable adherence to the reduction targets. Therefore, companies should not have to change the types of foods they offer in order to improve the nutritional quality of their menus, making the shift towards a healthier OOH sector a more achievable goal for industry.

We were not able to account for heterogeneity in item-level sales due to the known lack of accessible sales data from the OOH sector (26). It is possible that healthier menu items are responsible for a smaller proportion of sales, thus making little difference to diet-related health outcomes. Greater transparency from companies in regards to the proportion of their sales that come from healthy and less healthy foods would permit a more holistic analysis, and could be encouraged by governments mandating the reporting of this information.

The time and resource constraints of manually collecting data from individual websites limited our sample size of menu items, and meant that the completeness and accuracy of the data was largely dependent on restaurants’ own reporting. For example, we found instances of Toby Carvery underreporting kcal (detailed further in S2 Table), which we left as reported as we did not have the capacity to laboratory test all menu items to verify the information. Only five restaurants reported serving size for all of their menu items, therefore we often had to use subcategory average serving sizes to calculate per serving or per 100g information where missing, which may have differed to the true value given the variation we observed in nutritional content within subcategories. Not all menu items would have been included for each company, for example, we evaluated Burger King’s online menu rather than their more extensive ‘Nutrition Explorer’, due to the less complete nutritional information in the latter, and Subway now (2025) have ‘Cookies and Sweet Treats’ on their online menu which was not published during our data collection. These limitations reflect the lack of standardisation in reporting nutritional information across the OOH sector, highlighting another gap that government regulation could address.

We only assessed adherence at one time point, therefore we cannot determine which restaurants or subcategories have shown the most or least improvement in nutritional quality over time, which could provide further insight into which areas of the sector need more stringent monitoring.

### Comparison to Other Literature

To our knowledge, only two studies have looked at company-level changes in nutrient content in the UK, both focusing on the sugar reduction programme only, with one looking at manufacturers of grocery foods and the other at OOH. Bandy et al (19) found that in 2018, just under half (24/50) of the best-selling retailers manufacturers across five food categories (biscuits and cereal bars, breakfast cereals, chocolate confectionery, sugar confectionery, and yoghurts) met the intermediary 2018 sugar targets, and four companies had already met the 2020 targets. Our study found no companies met all of their sugar targets, potentially indicating greater engagement from the retailer grocery food sector compared to OOH. However, our study included a wider range of food categories, used nutritional data from 2024 (six years later), and Bandy et al used sales-weighted averages for sugar content (by brand sales volume), which we did not have the relevant data to replicate. Pepper et al (20) found that 4/48 OOH companies significantly reduced the average sugar per serving of their Desserts by at least 20%, from 2017 to 2020. Given that Bandy et al found four manufacturers had already met the 20% reduction target by 2018, this could provide another indication that the grocery sector is more engaged with the programme than OOH. Pepper et al observed large variation in sugar and calorie content between companies with different styles of food, but also between similar chains, which corroborates with findings from our study across all three target nutrients (sugar, salt, and calories) and beyond just Desserts.

Together, these findings suggest that whilst adherence to the targets may be low overall, there are examples of OOH companies that are performing well, and performance is not constrained by the type of cuisine being offered.

The most recent sugar reduction progress report found no OOH companies met the 20% reduction target by 2020 (12), and whilst this is consistent with our findings, it was based on data from only five companies, and four of which were sandwich/café restaurants (Costa, Pret, Greggs, and Starbucks). The most recent salt reduction progress report had insufficient data to present company-level performance for the OOH sector, although it reported that overall, 74% of OOH products met their salt target (26). Our study found only 58% of menu items met their salt target, potentially due to the progress report using the ‘maximum’ target values whilst we used the ‘average’ target values, which were less lenient (e.g., Dips had a ‘maximum’ target of 0.9g per 100g but an ‘average’ target of 0.75g). There is no relevant comparison between our findings and those in the calorie reduction progress report, as this report did not include any company-level analyses (13).

Reduction target programmes in other countries have been found to be effective in reducing the sugar, salt, calorie, and fat content of food products. One systematic review found that of 26 studies evaluating government-set reduction targets across 15 different countries, 22 found improvements in nutritional quality (27). However, the vast majority of these studies only focused on salt, and reported the percentage change in salt content over time rather than adherence to the targets specifically, so our findings may not be directly comparable. A study assessing OOH foods in the US found significantly fewer fast-food meals met the American Heart Association’s calorie guidelines in 2015, 2016, and 2017 compared to 2008, with no changes observed for saturated fat or sodium, suggesting that our findings are indicative of a wider global trend for poor nutritional quality in the OOH food sector. The restaurants included in our study are owned by multinational companies operating on a global scale, therefore our findings can provide insight into the nutritional quality of OOH foods beyond a UK context.

### Policy Implications

The overall low adherence to the targets could be a result of the programmes being voluntary, with research from other countries evidencing the increased effectiveness of mandatory versus voluntary nutrition policies in inciting reformulation (27). With no regular publication of progress reports to monitor companies’ adherence to the targets, there are minimal incentives for the companies to work towards them. More regular and granular reporting of adherence to the targets by individual restaurants in the OOH sector, as part of the Food Data Transparency Partnership for example (28), might lead to greater public scrutiny and thus greater engagement with the targets. Whilst the voluntary nature of the targets may reduce companies’ engagement, our study was able to demonstrate that restaurants with varying cuisines are able to make progress towards the targets, highlighting their attainability across the OOH sector.

The overall lower adherence to the sugar targets compared to the calorie and salt targets, could be a result of the deadline for the sugar targets being in 2020, versus 2024/5 for the calorie and salt targets. It is possible that adherence to the sugar targets has dropped since the deadline was reached, as there has been no subsequent revisions to the sugar targets to maintain food companies’ engagement with the programme. Alternatively, the sugar reduction targets were set per 100g, whilst the calorie and salt targets were primarily per serving (all calorie targets per serving, OOH-specific salt targets per serving), meaning progress towards the calorie and salt targets could be made through reducing menu items’ serving size, whilst the sugar targets could only be met through reformulation, which is potentially more time and resource intensive.

Unlike the retail sector, there is no mandatory reporting standard for nutritional information or serving size for the OOH sector, resulting in inconsistent and limited data being used to monitor adherence to the targets. Mandating standardised nutrition reporting would improve transparency in the OOH sector and make tracking compliance to the targets easier and more accurate, potentially inciting greater compliance from companies. Modelling work by Shangguan et al (29) found the drop from 100% to 50% industry compliance with US National Salt and Sugar Reduction targets approximately halved the averted cardiovascular disease events, QALYs gained, and net savings for healthcare observed over a lifetime, highlighting the importance of meeting the targets in full in order to bring about substantial benefits to public health.

Changes could be made to the targets themselves to incite greater engagement. All menu items had to be manually categorised into the relevant target categories to compare their nutrient content to the target value, which was time and resource intensive, and open to subjectivity and error. Using comparable categories across targets, or setting targets based on a single holistic measure of nutritional quality rather than individual nutrients (e.g., using the UK Nutrient Profiling Model), could remove some of the burden of self-monitoring placed onto companies, permitting greater progress.

In conclusion, our findings suggest there has been low engagement with the UK Government’s reduction targets from the OOH sector, which could suggest that mandatory regulations may be a more effective approach to improving the nutritional quality of OOH food. Whilst we found certain restaurant types to perform worse against the targets than others, restaurants offering similar menu items were also found to vary in target adherence, suggesting that companies should not have to change the focus of their menus in order to meet the targets, making them more attainable. This study highlights the need for accessible, standardised reporting of nutritional and serving size information from the OOH sector, to aid monitoring companies’ performance against the targets, and in turn inciting greater engagement from industry with the reduction programmes.

## Data Availability

The dataset used to conduct the analyses in this article has been deposited in the Oxford University Research Archive under CC-BY license (DOI: 10.5287/ora-6raddkrg9).

https://dx.doi.org/10.5287/ora-6raddkrg9

## Abbreviations

NCD: Non-communicable disease
OOH: Out of home

## Acknowledgements

There are no acknowledgements.

---

**S1 Table.** STROBE Checklist.

|  | Item No. | Recommendation | Respected? | Comments and quotes |
| --- | --- | --- | --- | --- |
| **Title and abstract** | 1 | (a) Indicate the study's design with a commonly used term in the title or the abstract | Yes | Study design is indicated in the title with the phrase 'A Cross-sectional Study' (beginning line 1) |
|  |  | (b) Provide in the abstract an informative and balanced summary of what was done and what was found | Yes | The abstract includes a 'Methods/Findings' section which provides information on our study approach and findings (beginning line 46) |
| **Introduction** |  |  |  |  |
| Background/rationale | 2 | Explain the scientific background and rationale for the investigation being reported | Yes | The Background section outlines the previous literature evaluating the UK Government's sugar, salt, and calorie reduction targets, followed up by how this study will address the outstanding gaps in this literature (beginning line 72). |
| Objectives | 3 | State specific objectives, including any prespecified hypotheses | Yes | The aim of this study is described at the end of the Background section: "This study aimed to assess adherence to the UK Government's sugar, salt, and calorie reduction targets for food items offered by the highest grossing restaurant chains in the UK, in 2024." (beginning line 135). No specific hypotheses were specified. |
| **Methods** |  |  |  |  |
| Study design | 4 | Present key elements of study design early in the paper | Yes | A summary of our methodology approach is given at the start of the methods section (beginning line 139). |
| Setting | 5 | Describe the setting, locations, and relevant dates, including periods of recruitment, exposure, follow-up, and data collection | Yes | As our study used nutritional data, there is no setting or location of the study. We do however include the months in which data collection took place (beginning line |
|  |  |  |  | 169): "Menu data was collected directly from restaurants' UK websites in February and March of 2024 (May 2024 for Subway)." |
| Participants | 6 | (a) Give the eligibility criteria, and the sources and methods of selection of participants | Yes | The 'Identifying Restaurants' subheading of the Methods section (beginning line 149) outlines how we selected the restaurants to include in our sample, and the criteria restaurants had to meet in order to be included. |
| Variables | 7 | Clearly define all outcomes, exposures, predictors, potential confounders, and effect modifiers. Give diagnostic criteria, if applicable | Yes | Our Analysis section outlines how we calculated our two outcomes of interest: average nutrient content (beginning line 241) and adherence to targets (beginning line 263). |
| Data sources/<br>measurement | 8* | For each variable of interest, give sources of data and details of methods of assessment (measurement). Describe comparability of assessment methods if there is more than one group | Yes | The 'Data Collection' subheading of the Methods section outlines the source of the nutritional data we collected (beginning line 168) |
| Bias | 9 | Describe any efforts to address potential sources of bias | Yes | N/A |
| Study size | 10 | Explain how the study size was arrived at | Yes | The 'Identifying Restaurants' subheading of the Methods section (beginning line 149) outlines the criteria for the restaurants we included in our sample, and the first paragraph of the 'Missing Data' subheading in the Analysis section outlines our exclusion criteria for menu items. |
| Quantitative variables | 11 | Explain how quantitative variables were handled in the analyses. If applicable, describe which groupings were chosen and why | Yes | Under their respective subheadings in the Analysis section, we describe how our two outcome variables (average nutrient content and target adherence) |
|  |  |  |  | were used for our analyses (beginning line 241 and 263). |
| Statistical methods | 12 | (a) Describe all statistical methods, including those used to control for confounding | Yes | The statistical analyses conducted for each of our two outcomes are described under their respective subheadings in the Analysis section (beginning line 241 and 263). |
|  |  | (b) Describe any methods used to examine subgroups and interactions | Yes | For some analyses, restaurants were assigned to one of five restaurant types. We describe the categorisation approach in the 'Categorising Menu Items' subheading of the Methods section (beginning line 187), and in the 'Adherence to Targets' subheading of the Analysis section we describe how these sub-groups were used when conducting our analyses (beginning line 280). |
|  |  | (c) Explain how missing data were addressed | Yes | The 'Missing Data' subheading of the Analysis section explains how we dealt with missing data (line beginning 218). |
|  |  | (d) If applicable, describe analytical methods taking account of sampling strategy |  | N/A |
|  |  | (e) Describe any sensitivity analyses | Yes | The sensitivity analysis we conducted is described under the 'Sensitivity Analysis' subheading of the Analysis section (beginning line 285). |
| **Results** |  |  |  |  |
| Participants | 13* | (a) Report numbers of individuals at each stage of study—eg numbers potentially eligible, examined for eligibility, confirmed eligible, included in the study, completing follow-up, and analysed | Yes | In the first paragraph of the Results section (beginning line 333), we provide the number of menu items excluded against each criterion, to go from our initially collected sample to our final sample of menu items. |
|  |  | (b) Give reasons for non-participation at each stage |  | N/A |
|  |  | (c) Consider use of a flow diagram | Yes | Use of a flow diagram was not deemed appropriate. |
| Descriptive data | 14* | (a) Give characteristics of study participants (eg demographic, clinical, social) and information on exposures and potential confounders | Yes | Table 1 (beginning line 344) provides an overview of the characteristics for each restaurant, including Brand Name, Company Name, Restaurant Type, 2022 Sales Value, Number of Menu Items Included, and Number of Unique Subcategories Included. S2 Table also provides further details of the data collected for each restaurant (beginning line 821). |
|  |  | (b) Indicate number of participants with missing data for each variable of interest | Yes | In the first paragraph of the Results section (beginning line 333), we provide the number of menu items excluded against each criterion, to go from our initially collected sample to our final sample of menu items. |
| Outcome data | 15* | Report numbers of outcome events or summary measures | Yes | All outcomes by restaurant and subcategory are reported in supplementary tables. |
| Main results | 16 | (a) Give unadjusted estimates and, if applicable, confounder-adjusted estimates and their precision (eg, 95% confidence interval). Make clear which confounders were adjusted for and why they were included | Yes | Full outcomes for all statistical analyses are provided in supplementary tables. For Dunn tests, the supplementary tables provide *p* values and alpha level with Bonferroni correction, and for logistic regressions, the supplementary tables provide *p* values, alpha level with Bonferroni correction, odds ratios, and confidence intervals for odds ratios. In their respective subheadings under the Analysis section, it is explained how Bonferroni corrections were applied (beginning line 256 for 'average nutrient content' analyses and line 280 for 'adherence to targets' analyses). |
|  |  | (b) Report category boundaries when continuous variables were categorized |  | N/A |
|  |  | (c) If relevant, consider translating estimates of relative risk into absolute risk for a meaningful time period |  | N/A |
| Other analyses | 17 | Report other analyses done—eg analyses of subgroups and interactions, and sensitivity analyses | Yes | Findings from the sensitivity analysis are reported from line 564, and full outcomes are reported in supplementary tables S21-22 (beginning line 971). |
| **Discussion** |  |  |  |  |
| Key results | 18 | Summarise key results with reference to study objectives | Yes | Key findings are summarised in the first paragraph of the discussion (beginning line 577) |
| Limitations | 19 | Discuss limitations of the study, taking into account sources of potential bias or imprecision. Discuss both direction and magnitude of any potential bias | Yes | Limitations of the study are described in the 'Strengths and Limitations' section of the discussion (beginning line 592). |
| Interpretation | 20 | Give a cautious overall interpretation of results considering objectives, limitations, multiplicity of analyses, results from similar studies, and other relevant evidence | Yes | We make considerations for how the limitations of the study may have impacted our findings in the 'Strengths and Limitations' section (beginning line 592), and interpret our results within the context of the wider literature in the 'Comparison to Other Literature' section (beginning line 626) |
| Generalisability | 21 | Discuss the generalisability (external validity) of the study results | Yes | We describe how our findings are generalisable beyond a UK context (line beginning 664). |
| **Other information** |  |  |  |  |
| Funding | 22 | Give the source of funding and the role of the funders for the present study and, if applicable, for the original study on which the present article is based | Yes | Funding sources and role of funders in this study is provided on the title page (beginning line 12). |

**S2 Table.** Overview of data collection approach and completeness of collected data, for each restaurant. Restaurants in descending order by number of menu items.

| Restaurant | No. of Menu Items | Per 100g or Per Serving Information? | Serving Size (g) Provided? | Data Collection Method | Missing Nutrition Information | Notes |
| --- | --- | --- | --- | --- | --- | --- |
| **Pizza Hut** | 330 | Majority per serving (per 100g only for other brand ice-creams) | Only for four products | Manual input from PDF | Kilojoules, Sodium, Fibre |  |
| **Hungry Horse** | 247 | Per serving | No | PDF | Sodium, Fibre |  |
| **Pizza Express** | 247 | Majority both (15 items with per 100g only) | No | PDF | Sodium |  |
| **Domino's** | 241 | Majority both (per 100g only for other brand ice-creams) | No | PDF |  |  |
| **Prezzo** | 218 | Per serving | No | PDF | Calories (had to calculate from fat, carbohydrates, and protein), Sodium |  |
| **Toby Carvery** | 214 | Per serving | No | Web-scrape | Sodium, Fibre | When comparing kcal as reported to kcal as calculated from protein, fat, and carbohydrates, 17 products had a reported kcal that was at least 100kcal lower than the calculated kcal (ranging from a 114.68 to 936.98 difference), suggesting some inaccuracy in their reporting. |
| **Papa John's** | 180 | Majority both (37 items per 100g only) | Yes | Manual input from PDF |  |  |
| **Harvester** | 175 | Per serving | For 13 items | PDF | Sodium, Fibre |  |
| **Greggs** | 174 | Both | Yes | PDF | Sodium |  |
| **Pret** | 151 | Majority both (12 items with per 100g only) | No | Web-scrape | Sodium |  |
| **Costa** | 123 | Both | Yes | PDF | Sodium, some Fibre (only provided for other brand products) |  |
| **McDonald's** | 111 | Per serving | No | Web-scrape | Sodium |  |
| **Caffé Nero** | 108 | Both | Yes | PDF | Sodium, some Fibre |  |
| **Wagamama** | 105 | Both | No | Web-scrape |  |  |
| **Nando's** | 90 | Per serving | No | Web-scrape | Sodium |  |
| **Subway** | 80 | Per serving | Yes | Web-scrape | Sodium |  |
| **KFC** | 76 | Per serving | No | Web-scrape | Sodium, Fibre |  |
| **Vintage Inns** | 73 | Per serving | No | PDF | Sodium, Fibre |  |
| **Leon** | 67 | Per serving | Yes | Manual input from website | Kilojoules, Sodium |  |
| **Starbucks** | 49 | Per serving | No | PDF | Sodium |  |
| **Burger King** | 40 | Majority both (1 item per 100g only) | No | Manual input from website | Sodium, Fibre | In addition to their menu webpage, Burger King had a 'Nutrition Explorer' website. Nutritional information was taken from the menu webpage as this provided both per 100g and per serving information, whereas the Nutrition Explorer only provided per serving information. |

**S3 Table.** Categorisation criteria for the 12 broad food subcategories.

| Category | Subcategory | Definition |
| --- | --- | --- |
| **Main meals** | Burgers | Burgers of all sizes |
|  | Chicken | All main chicken dishes. Chicken products described as a 'side' to be included under 'other sides'. |
|  | Pizzas | Pizzas of all sizes. |
|  | Breakfast items | Any items described as breakfast items, such as bacon and egg sandwiches. |
|  | Children's meals | Any main meal described as a kid's meal. Main meals only, other products (e.g., sides) to be included in other categories accordingly. |
|  | Sandwiches | All products described as a sandwich, wrap or slice, including hot sandwiches, paninis, pasties and sausage rolls. |
|  | Salads | All salads as a main item. Side salad and green salads to be included under 'other sides'. |
|  | Other mains | All other mains, including curries, fish and chips, roasts, and steaks. |
| **Sides/Extras** | Potato sides | All fries, chips, wedges and other potato side dishes |
|  | Other sides | All other products described as a 'side', including side salads, soups and side portions of chicken etc |
|  | Sauces | All sauces and condiments |
| **Desserts** | Desserts | Includes all desserts, cakes, pastries, ice creams and sweet treats |

**S4 Table.** The range of calorie, salt (per 100g and per serving), and sugar target values set for menu items within each subcategory.

| Subcategory | Calorie Target Values<br>(per serving) | Salt Target Values (per<br>100g) | Salt Target Values (per<br>serving) | Sugar Target Values<br>(per 100g) |
| --- | --- | --- | --- | --- |
| Breakfast Items | 485 | 2.11 | 2.70 | 28.70 |
| Burgers | 380 | 0.00 | 2.20 |  |
| Chicken | 0 |  | 3.15 |  |
| Children's Meals | 485 |  | 0.09 | 0.00 |
| Desserts |  | 1.07 |  | 38.40 |
| Other Mains | 485 | 0.80 | 2.25 |  |
| Other Sides | 745 | 2.16 | 3.15 | 0.00 |
| Pizzas | 0 |  | 5.12 |  |
| Potato Sides | 260 | 0.53 | 0.47 |  |
| Salads | 485 |  |  |  |
| Sandwiches | 50 |  | 1.58 |  |
| Sauces |  | 2.13 |  |  |

**S5 Table.** The mean, median, and standard deviation, for Kcal per 100g, per recommended serving, and per subcategory average serving, across all menu items in each subcategory. In descending order by Mean Kcal per 100g.

| Subcategory | Per 100g |  |  | Per Reported Serving |  |  | Per Subcategory Average Serving |  |  |
| --- | --- | --- | --- | --- | --- | --- | --- | --- | --- |
|  | Mean | Median | SD | Mean | Median | SD | Mean | Median | SD |
| **Dessert** | 409.98 | 390.00 | 276.53 | 314.15 | 280.00 | 248.05 | 319.49 | 303.92 | 215.49 |
| **Potato Sides** | 315.73 | 244.90 | 261.39 | 323.28 | 269.75 | 284.12 | 353.09 | 273.88 | 292.31 |
| **Pizzas** | 313.24 | 288.02 | 88.79 | 668.08 | 695.55 | 251.39 | 638.37 | 586.96 | 180.95 |
| **Burgers** | 308.36 | 256.60 | 193.85 | 737.24 | 603.60 | 471.38 | 738.18 | 614.27 | 464.06 |
| **Sauces** | 298.44 | 202.99 | 305.45 | 92.33 | 62.00 | 97.58 | 91.15 | 62.00 | 93.29 |
| **Chicken** | 280.77 | 212.87 | 197.96 | 680.66 | 568.00 | 487.79 | 683.87 | 518.50 | 482.17 |
| **Sandwiches** | 250.45 | 232.37 | 87.87 | 474.07 | 449.20 | 188.71 | 476.93 | 442.50 | 167.32 |
| **Other Sides** | 233.80 | 209.92 | 190.10 | 259.64 | 225.75 | 201.30 | 267.30 | 240.00 | 217.34 |
| **Breakfast Items** | 230.83 | 200.12 | 193.27 | 375.49 | 316.00 | 325.61 | 389.87 | 338.00 | 326.43 |
| **Children's Meals** | 196.37 | 175.49 | 96.53 | 371.59 | 343.50 | 188.22 | 388.33 | 347.03 | 190.89 |
| **Other Mains** | 163.43 | 156.20 | 79.63 | 755.97 | 738.00 | 387.47 | 769.51 | 735.50 | 374.96 |
| **Salads** | 145.91 | 134.50 | 79.66 | 334.34 | 310.00 | 210.58 | 320.33 | 295.28 | 174.88 |

**S6 Table.** The mean, median, and standard deviation, for Salt per 100g, per recommended serving, and per subcategory average serving, across all menu items in each subcategory. In descending order by Mean Salt per 100g.

| Subcategory | Per 100g |  |  | Per Reported Serving |  |  | Per Subcategory Average Serving |  |  |
| --- | --- | --- | --- | --- | --- | --- | --- | --- | --- |
|  | Mean | Median | SD | Mean | Median | SD | Mean | Median | SD |
| **Sauces** | 2.19 | 1.47 | 2.44 | 0.66 | 0.44 | 0.73 | 0.67 | 0.45 | 0.74 |
| **Pizza** | 1.59 | 1.43 | 0.65 | 3.44 | 3.26 | 1.74 | 3.25 | 2.92 | 1.32 |
| **Other Side** | 1.33 | 1.00 | 1.98 | 1.48 | 1.04 | 2.24 | 1.52 | 1.14 | 2.23 |
| **Burger** | 1.18 | 1.10 | 0.56 | 2.83 | 2.64 | 1.37 | 2.83 | 2.63 | 1.33 |
| **Sandwich** | 1.12 | 1.00 | 0.45 | 2.12 | 1.89 | 0.96 | 2.12 | 1.90 | 0.86 |
| **Chicken** | 1.08 | 0.84 | 0.85 | 2.63 | 2.10 | 2.10 | 2.64 | 2.05 | 2.08 |
| **Breakfast Item** | 0.98 | 0.77 | 1.12 | 1.64 | 1.11 | 1.91 | 1.66 | 1.29 | 1.89 |
| **Children's Meal** | 0.88 | 0.79 | 0.54 | 1.66 | 1.48 | 1.04 | 1.74 | 1.57 | 1.06 |
| **Potato Side** | 0.76 | 0.72 | 0.51 | 0.81 | 0.67 | 0.62 | 0.85 | 0.80 | 0.57 |
| **Salad** | 0.75 | 0.66 | 0.43 | 1.63 | 1.50 | 1.00 | 1.64 | 1.45 | 0.93 |
| **Other Main** | 0.74 | 0.70 | 0.41 | 3.44 | 3.09 | 1.97 | 3.50 | 3.29 | 1.92 |
| **Dessert** | 0.48 | 0.36 | 0.69 | 0.39 | 0.26 | 0.66 | 0.37 | 0.28 | 0.54 |

**S7 Table.** The mean, median, and standard deviation, for Sugar per 100g, per recommended serving, and per subcategory average serving, across all menu items in each subcategory. In descending order by Mean Sugar per 100g.

| Subcategory | Per 100g |  |  | Per Reported Serving |  |  | Per Subcategory Average Serving |  |  |
| --- | --- | --- | --- | --- | --- | --- | --- | --- | --- |
|  | Mean | Median | SD | Mean | Median | SD | Mean | Median | SD |
| Dessert | 34.28 | 28.23 | 26.52 | 26.18 | 20.66 | 22.90 | 26.72 | 22.00 | 20.66 |
| Sauces | 15.29 | 8.04 | 15.98 | 4.46 | 2.40 | 4.78 | 4.67 | 2.46 | 4.88 |
| Breakfast Item | 6.72 | 3.70 | 8.86 | 10.32 | 6.20 | 12.60 | 11.35 | 6.25 | 14.97 |
| Pizza | 4.73 | 4.28 | 2.27 | 10.11 | 8.00 | 6.38 | 9.63 | 8.73 | 4.62 |
| Burger | 4.25 | 3.93 | 2.70 | 10.15 | 9.20 | 6.52 | 10.18 | 9.40 | 6.47 |
| Other Side | 3.96 | 2.20 | 5.31 | 4.34 | 2.40 | 5.48 | 4.53 | 2.52 | 6.07 |
| Chicken | 3.96 | 1.04 | 5.37 | 9.71 | 1.79 | 13.08 | 9.65 | 2.53 | 13.09 |
| Sandwich | 3.65 | 3.31 | 2.41 | 7.14 | 5.94 | 5.17 | 6.95 | 6.30 | 4.59 |
| Children's Meal | 3.17 | 2.48 | 3.00 | 6.10 | 4.75 | 5.97 | 6.27 | 4.90 | 5.93 |
| Salad | 2.62 | 2.28 | 1.28 | 6.01 | 4.90 | 4.04 | 5.75 | 4.99 | 2.81 |
| Other Main | 2.58 | 2.04 | 2.03 | 12.05 | 9.50 | 9.73 | 12.13 | 9.60 | 9.54 |
| Potato Side | 2.07 | 1.23 | 3.51 | 2.21 | 1.30 | 3.93 | 2.31 | 1.37 | 3.92 |

**S8 Table.** The mean, median, and standard deviation, for Fat per 100g, per recommended serving, and per subcategory average serving, across all menu items in each subcategory. In descending order by Mean Fat per 100g.

| Subcategory | Per 100g |  |  | Per Reported Serving |  |  | Per Subcategory Average Serving |  |  |
| --- | --- | --- | --- | --- | --- | --- | --- | --- | --- |
|  | Mean | Median | SD | Mean | Median | SD | Mean | Median | SD |
| **Sauces** | 21.81 | 6.22 | 32.92 | 6.80 | 1.90 | 10.14 | 6.66 | 1.90 | 10.06 |
| **Dessert** | 19.61 | 17.40 | 16.34 | 14.92 | 12.99 | 13.30 | 15.28 | 13.56 | 12.74 |
| **Burger** | 15.73 | 12.85 | 11.84 | 37.74 | 29.15 | 28.84 | 37.65 | 30.75 | 28.34 |
| **Potato Side** | 14.09 | 10.77 | 10.40 | 13.96 | 12.00 | 10.12 | 15.76 | 12.04 | 11.63 |
| **Other Side** | 12.84 | 10.50 | 14.22 | 14.21 | 10.65 | 15.61 | 14.68 | 12.00 | 16.25 |
| **Chicken** | 12.82 | 9.48 | 8.90 | 31.05 | 23.60 | 21.90 | 31.22 | 23.10 | 21.68 |
| **Pizza** | 11.76 | 10.30 | 5.49 | 25.04 | 23.87 | 12.97 | 23.96 | 21.00 | 11.18 |
| **Sandwich** | 10.61 | 9.12 | 6.54 | 20.02 | 17.94 | 12.60 | 20.21 | 17.36 | 12.45 |
| **Breakfast Item** | 9.80 | 7.90 | 10.78 | 16.51 | 12.00 | 18.26 | 16.55 | 13.34 | 18.21 |
| **Salad** | 8.27 | 7.45 | 5.50 | 19.30 | 17.30 | 14.11 | 18.17 | 16.36 | 12.08 |
| **Other Main** | 7.77 | 6.58 | 4.80 | 35.47 | 32.05 | 22.04 | 36.59 | 31.00 | 22.59 |
| **Children's Meal** | 7.33 | 6.07 | 5.51 | 13.97 | 12.00 | 10.77 | 14.50 | 12.00 | 10.89 |

**S9 Table.**
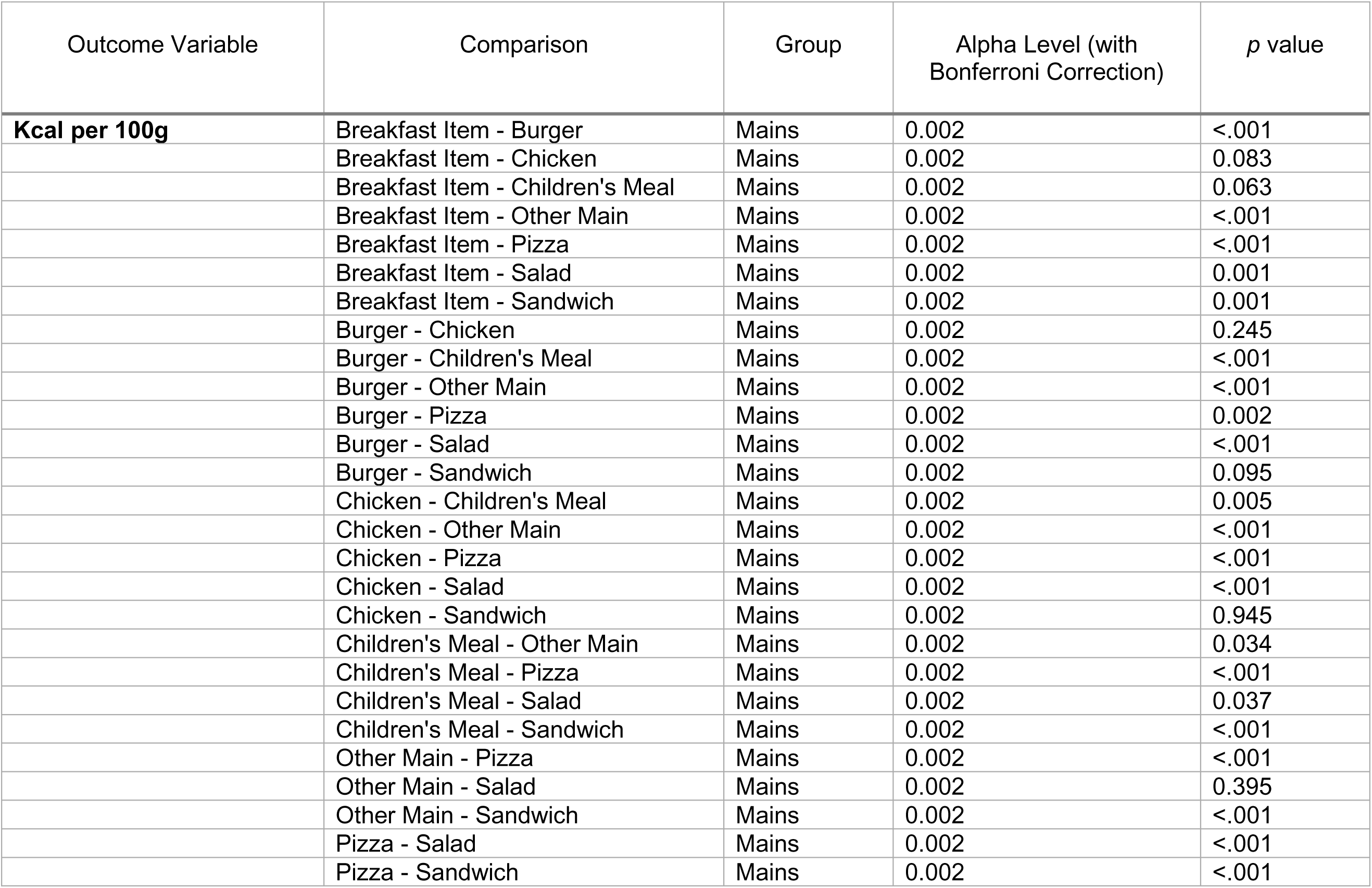
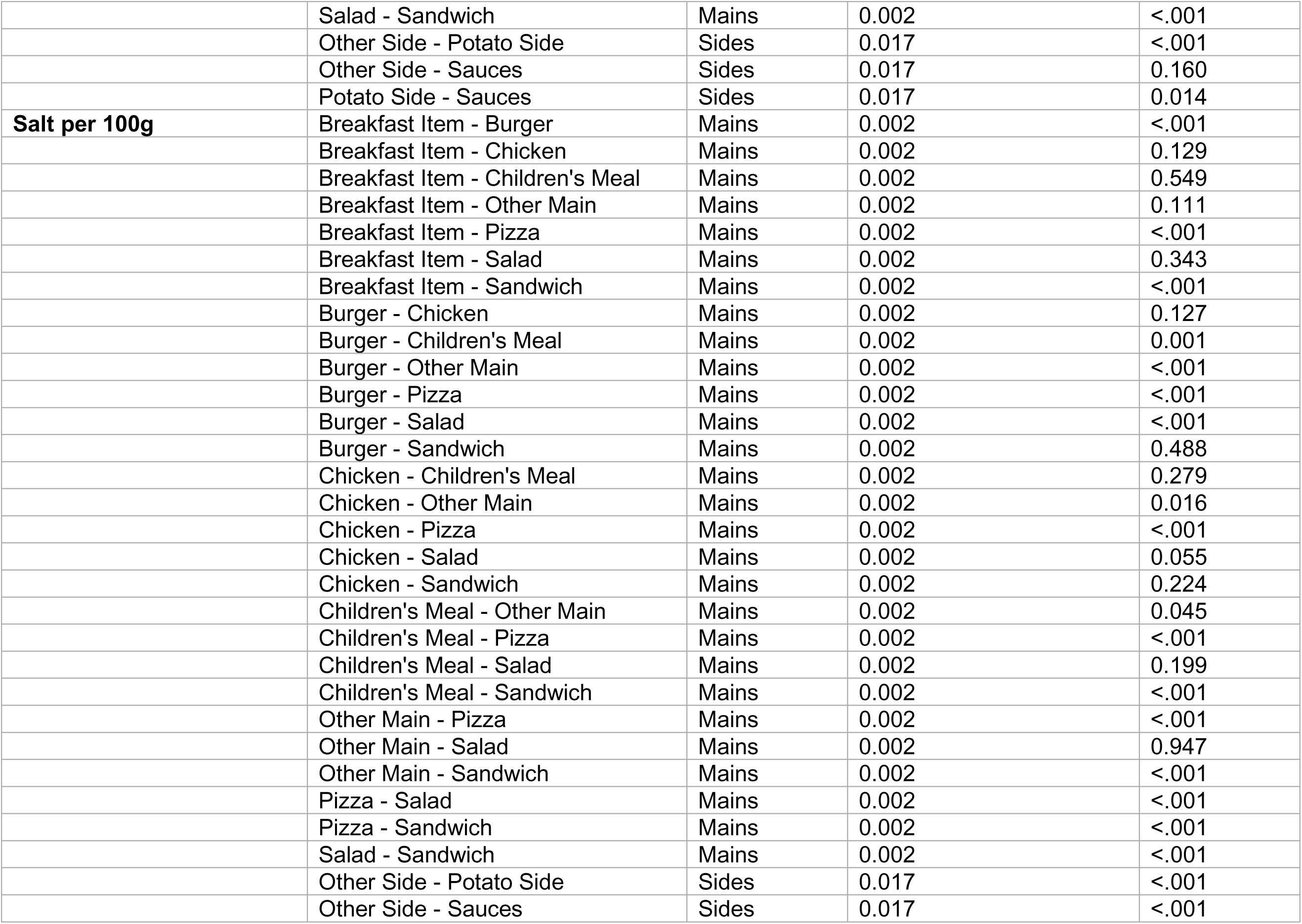
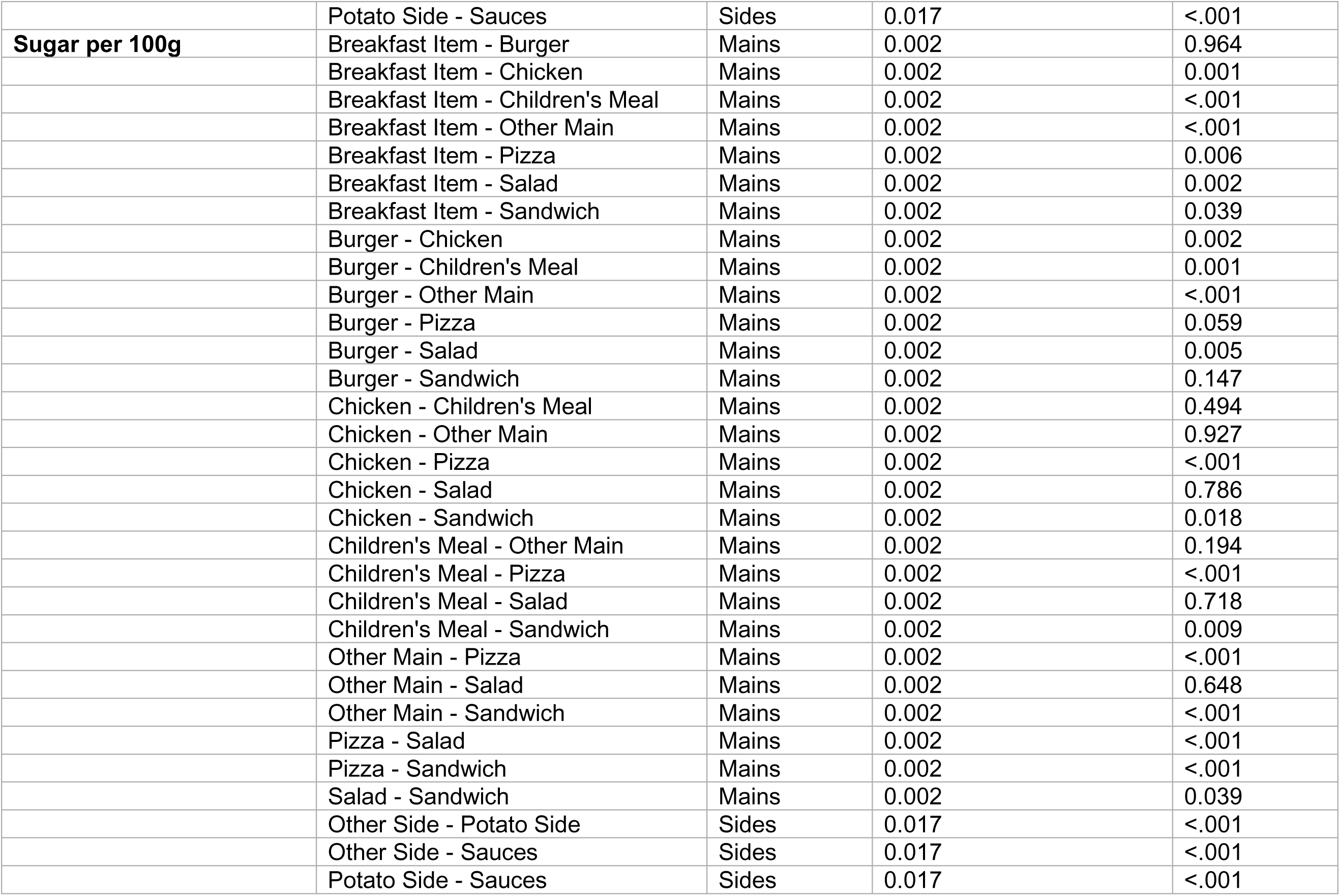
The alpha level and p value for pairwise comparisons between subcategories in kcal, sugar, and salt per 100g.

**S10 Table.** The mean, median, and standard deviation, for Kcal per 100g, per recommended serving, and per subcategory average serving, across all menu items in each restaurant. In descending order by Mean Kcal per 100g.

| Restaurant | Per 100g |  |  | Per Reported Serving |  |  | Per Subcategory Average Serving |  |  |
| --- | --- | --- | --- | --- | --- | --- | --- | --- | --- |
|  | Mean | Median | SD | Mean | Median | SD | Mean | Median | SD |
| **Caffé Nero** | 367.85 | 374.00 | 129.51 | 306.10 | 314.93 | 132.20 | 388.81 | 360.03 | 136.94 |
| **Vintage Inns** | 360.24 | 225.85 | 335.69 | 563.53 | 423.50 | 358.25 | 563.53 | 423.50 | 358.25 |
| **Costa** | 355.23 | 373.00 | 121.84 | 288.37 | 316.00 | 154.60 | 361.91 | 365.49 | 130.01 |
| **Prezzo** | 340.85 | 268.89 | 270.06 | 644.38 | 636.50 | 410.98 | 644.38 | 636.50 | 410.98 |
| **Pizza Hut** | 334.64 | 335.88 | 88.27 | 641.39 | 673.33 | 227.39 | 640.04 | 673.33 | 229.90 |
| **Harvester** | 317.08 | 239.65 | 265.08 | 527.68 | 346.00 | 470.09 | 548.03 | 367.00 | 462.94 |
| **Hungry Horse** | 310.72 | 237.91 | 325.23 | 555.58 | 382.00 | 520.09 | 555.58 | 382.00 | 520.09 |
| **Starbucks** | 301.27 | 251.63 | 167.78 | 305.16 | 301.00 | 126.76 | 305.16 | 301.00 | 126.76 |
| **Nando's** | 291.42 | 219.58 | 313.71 | 364.71 | 301.20 | 229.61 | 389.56 | 301.20 | 361.78 |
| **Greggs** | 271.75 | 259.00 | 106.12 | 351.33 | 348.04 | 179.16 | 391.68 | 382.24 | 189.30 |
| **Domino's** | 265.75 | 266.00 | 67.44 | 546.57 | 615.00 | 224.33 | 460.84 | 507.45 | 160.09 |
| **Pret** | 262.78 | 238.00 | 139.01 | 344.88 | 347.00 | 158.07 | 394.39 | 388.47 | 141.67 |
| **Papa John's** | 260.19 | 255.00 | 65.19 | 295.72 | 240.69 | 211.54 | 439.71 | 490.13 | 147.87 |
| **Burger King** | 252.75 | 256.10 | 29.33 | 459.85 | 426.00 | 229.95 | 477.07 | 524.43 | 184.25 |
| **Toby Carvery** | 248.43 | 138.39 | 299.44 | 291.89 | 195.00 | 329.29 | 304.69 | 195.00 | 354.43 |
| **Pizza Express** | 224.13 | 213.00 | 105.96 | 473.13 | 391.00 | 337.93 | 330.66 | 355.57 | 168.69 |
| **Leon** | 223.03 | 184.15 | 117.02 | 357.00 | 334.00 | 173.91 | 353.22 | 340.05 | 191.23 |
| **KFC** | 206.24 | 181.66 | 116.47 | 291.05 | 282.50 | 203.07 | 291.05 | 282.50 | 203.07 |
| **Subway** | 196.55 | 209.48 | 71.65 | 325.69 | 311.50 | 155.74 | 343.47 | 377.25 | 120.09 |
| **McDonald's** | 194.11 | 182.97 | 108.12 | 308.46 | 319.00 | 175.36 | 308.46 | 319.00 | 175.36 |
| **Wagamama** | 161.78 | 152.00 | 78.59 | 520.69 | 451.00 | 327.31 | 405.15 | 346.06 | 242.85 |

**S11 Table.** The mean, median, and standard deviation, for Salt per 100g, per recommended serving, and per subcategory average serving, across all menu items in each restaurant. In descending order by Mean Salt per 100g.

| Restaurant | Per 100g |  |  | Per Reported Serving |  |  | Per Subcategory Average Serving |  |  |
| --- | --- | --- | --- | --- | --- | --- | --- | --- | --- |
|  | Mean | Median | SD | Mean | Median | SD | Mean | Median | SD |
| **Prezzo** | 1.87 | 1.34 | 1.64 | 3.67 | 3.60 | 2.69 | 3.67 | 3.60 | 2.69 |
| **Papa John's** | 1.61 | 1.38 | 2.77 | 1.78 | 1.29 | 3.25 | 2.61 | 2.55 | 3.16 |
| **Pizza Hut** | 1.53 | 1.52 | 0.52 | 2.93 | 2.96 | 1.15 | 2.92 | 2.96 | 1.16 |
| **Domino's** | 1.41 | 1.32 | 0.84 | 2.91 | 2.98 | 1.53 | 2.44 | 2.55 | 1.13 |
| **Hungry Horse** | 1.21 | 0.78 | 1.56 | 2.02 | 1.28 | 2.15 | 2.02 | 1.28 | 2.15 |
| **Pizza Express** | 1.16 | 1.10 | 1.05 | 2.63 | 2.08 | 2.29 | 1.84 | 1.94 | 1.50 |
| **Nando's** | 1.02 | 0.77 | 1.18 | 1.29 | 1.30 | 0.91 | 1.38 | 1.30 | 1.35 |
| **Burger King** | 1.02 | 1.10 | 0.43 | 1.92 | 1.84 | 1.23 | 2.02 | 2.11 | 1.12 |
| **KFC** | 0.98 | 0.72 | 1.12 | 1.38 | 0.92 | 1.25 | 1.38 | 0.92 | 1.25 |
| **Greggs** | 0.95 | 0.83 | 0.63 | 1.24 | 1.30 | 0.80 | 1.47 | 1.47 | 0.99 |
| **Subway** | 0.95 | 0.95 | 0.37 | 1.58 | 1.50 | 0.79 | 1.69 | 1.75 | 0.69 |
| **Harvester** | 0.93 | 0.72 | 0.95 | 1.72 | 1.05 | 2.09 | 1.76 | 1.06 | 2.08 |
| **Vintage Inns** | 0.91 | 0.66 | 0.82 | 1.85 | 1.63 | 1.37 | 1.85 | 1.63 | 1.37 |
| **Pret** | 0.82 | 0.78 | 0.59 | 1.39 | 1.41 | 1.07 | 1.58 | 1.53 | 1.23 |
| **Toby Carvery** | 0.81 | 0.39 | 1.38 | 1.02 | 0.50 | 1.47 | 1.10 | 0.51 | 1.80 |
| **Leon** | 0.78 | 0.72 | 0.50 | 1.38 | 1.30 | 1.05 | 1.41 | 1.29 | 1.20 |
| **Wagamama** | 0.75 | 0.62 | 0.65 | 2.54 | 2.28 | 2.18 | 2.00 | 1.75 | 1.53 |
| **McDonald's** | 0.73 | 0.58 | 0.52 | 1.21 | 1.10 | 0.94 | 1.21 | 1.10 | 0.94 |
| **Costa** | 0.62 | 0.55 | 0.43 | 0.63 | 0.38 | 0.63 | 0.76 | 0.44 | 0.71 |
| **Caffé Nero** | 0.61 | 0.52 | 0.46 | 0.71 | 0.46 | 0.75 | 0.86 | 0.46 | 0.88 |
| **Starbucks** | 0.54 | 0.47 | 0.39 | 0.67 | 0.40 | 0.67 | 0.67 | 0.40 | 0.67 |

**S12 Table.** The mean, median, and standard deviation, for Sugar per 100g, per recommended serving, and per subcategory average serving, across all menu items in each restaurant. In descending order by Mean Sugar per 100g.

| Restaurant | Per 100g |  |  | Per Reported Serving |  |  | Per Subcategory Average Serving |  |  |
| --- | --- | --- | --- | --- | --- | --- | --- | --- | --- |
|  | Mean | Median | SD | Mean | Median | SD | Mean | Median | SD |
| **Costa** | 21.99 | 23.10 | 17.48 | 13.52 | 7.64 | 12.29 | 18.28 | 18.70 | 13.06 |
| **Caffé Nero** | 20.68 | 20.80 | 17.45 | 14.63 | 11.88 | 11.35 | 17.77 | 18.01 | 12.77 |
| **Vintage Inns** | 15.80 | 3.31 | 29.26 | 16.33 | 5.80 | 22.17 | 16.33 | 5.80 | 22.17 |
| **Harvester** | 15.33 | 5.76 | 26.20 | 17.23 | 8.30 | 22.29 | 18.60 | 9.10 | 23.57 |
| **Starbucks** | 15.27 | 12.06 | 13.91 | 13.37 | 12.00 | 10.04 | 13.37 | 12.00 | 10.04 |
| **Toby Carvery** | 12.51 | 3.85 | 19.45 | 11.45 | 4.85 | 15.15 | 11.65 | 5.00 | 15.18 |
| **Hungry Horse** | 10.53 | 3.27 | 29.29 | 12.50 | 5.90 | 23.51 | 12.50 | 5.90 | 23.51 |
| **Greggs** | 10.40 | 3.80 | 13.01 | 10.66 | 6.64 | 13.47 | 10.25 | 7.02 | 9.13 |
| **Pret** | 9.52 | 3.40 | 12.57 | 9.87 | 6.22 | 9.06 | 10.90 | 7.09 | 10.24 |
| **Nando's** | 9.46 | 2.14 | 16.39 | 8.60 | 4.20 | 12.41 | 8.76 | 4.20 | 12.46 |
| **McDonald's** | 9.32 | 3.59 | 12.84 | 10.08 | 7.30 | 9.78 | 10.08 | 7.30 | 9.78 |
| **KFC** | 9.02 | 4.23 | 13.27 | 8.88 | 6.45 | 9.67 | 8.88 | 6.45 | 9.67 |
| **Leon** | 7.21 | 3.09 | 9.66 | 8.16 | 6.00 | 7.03 | 7.97 | 5.97 | 6.97 |
| **Prezzo** | 7.11 | 2.76 | 13.22 | 10.05 | 7.10 | 10.83 | 10.05 | 7.10 | 10.83 |
| **Domino's** | 6.65 | 6.00 | 7.06 | 12.41 | 14.16 | 7.75 | 10.49 | 12.02 | 6.44 |
| **Pizza Express** | 6.51 | 3.10 | 10.21 | 10.00 | 5.77 | 11.46 | 7.55 | 5.09 | 8.43 |
| **Papa John's** | 6.09 | 5.00 | 6.21 | 7.19 | 4.55 | 14.02 | 9.22 | 9.99 | 5.34 |
| **Burger King** | 5.30 | 3.70 | 6.45 | 8.01 | 7.09 | 6.13 | 8.20 | 7.78 | 5.41 |
| **Wagamama** | 5.19 | 3.09 | 5.70 | 12.52 | 10.56 | 9.24 | 9.71 | 8.07 | 6.11 |
| **Pizza Hut** | 4.59 | 3.27 | 5.47 | 7.50 | 6.60 | 4.64 | 7.49 | 6.60 | 4.65 |
| **Subway** | 3.37 | 3.59 | 1.62 | 5.93 | 5.10 | 4.21 | 6.19 | 6.66 | 3.14 |

**S13 Table.** The mean, median, and standard deviation, for Fat per 100g, per recommended serving, and per subcategory average serving, across all menu items in each restaurant. In descending order by Mean Fat per 100g.

|  | Per 100g |  |  | Per Reported Serving |  |  | Per Subcategory Average Serving |  |  |
| --- | --- | --- | --- | --- | --- | --- | --- | --- | --- |
| Restaurant | Mean | Median | SD | Mean | Median | SD | Mean | Median | SD |
| Vintage Inns | 19.35 | 11.79 | 20.23 | 29.91 | 24.90 | 20.31 | 29.91 | 24.90 | 20.31 |
| Prezzo | 19.16 | 13.42 | 23.29 | 32.58 | 29.00 | 24.56 | 32.58 | 29.00 | 24.56 |
| Caffé Nero | 18.71 | 17.95 | 12.69 | 14.80 | 14.63 | 7.45 | 19.60 | 17.82 | 12.58 |
| Costa | 15.49 | 15.00 | 9.60 | 12.53 | 13.05 | 8.34 | 15.43 | 15.03 | 9.04 |
| Harvester | 14.98 | 11.28 | 14.88 | 26.19 | 16.70 | 25.94 | 26.40 | 17.00 | 25.81 |
| Hungry Horse | 14.39 | 9.24 | 17.37 | 25.40 | 16.20 | 26.65 | 25.40 | 16.20 | 26.65 |
| Nando's | 14.06 | 10.10 | 16.30 | 16.70 | 15.35 | 12.30 | 17.69 | 15.35 | 16.50 |
| Starbucks | 13.97 | 11.37 | 9.00 | 13.51 | 14.00 | 6.34 | 13.51 | 14.00 | 6.34 |
| Toby Carvery | 13.40 | 6.24 | 19.40 | 16.42 | 8.00 | 22.45 | 17.24 | 8.00 | 23.82 |
| Pret | 13.05 | 11.20 | 9.40 | 16.72 | 14.97 | 9.56 | 19.37 | 18.63 | 10.06 |
| Pizza Hut | 13.01 | 12.25 | 4.61 | 24.46 | 22.33 | 9.88 | 24.40 | 22.33 | 9.97 |
| Burger King | 12.93 | 12.70 | 2.80 | 24.06 | 20.78 | 14.36 | 24.60 | 26.57 | 10.90 |
| Leon | 11.70 | 9.43 | 8.64 | 17.39 | 16.00 | 9.52 | 17.31 | 17.39 | 11.37 |
| Greggs | 11.62 | 9.55 | 8.29 | 14.03 | 12.58 | 9.67 | 16.48 | 14.66 | 12.88 |
| Domino's | 11.01 | 10.50 | 7.37 | 20.98 | 19.22 | 8.89 | 17.83 | 16.91 | 6.36 |
| Papa John's | 10.83 | 9.25 | 6.40 | 12.51 | 8.81 | 11.61 | 17.25 | 16.10 | 8.08 |
| Pizza Express | 10.70 | 7.40 | 11.14 | 18.63 | 15.09 | 14.00 | 13.50 | 12.84 | 9.02 |
| KFC | 9.10 | 7.90 | 7.99 | 12.41 | 10.95 | 9.65 | 12.41 | 10.95 | 9.65 |
| McDonald's | 7.82 | 6.51 | 6.80 | 12.41 | 11.30 | 9.99 | 12.41 | 11.30 | 9.99 |
| Subway | 7.68 | 7.63 | 4.57 | 13.02 | 13.00 | 8.71 | 13.03 | 13.36 | 7.06 |
| Wagamama | 7.39 | 6.14 | 5.79 | 22.25 | 19.06 | 15.64 | 17.50 | 15.72 | 12.07 |

**S14 Table.** The mean, median, and standard deviation, for Kcal per 100g, per recommended serving, and per subcategory average serving, across all menu items in each restaurant group. In descending order by Mean Kcal per 100g.

| Restaurant Type | Per 100g |  |  | Per Reported Serving |  |  | Per Subcategory Average Serving |  |  |
| --- | --- | --- | --- | --- | --- | --- | --- | --- | --- |
|  | Mean | Median | SD | Mean | Median | SD | Mean | Median | SD |
| **Sandwich** | 293.24 | 267.07 | 133.85 | 325.17 | 331.00 | 158.47 | 374.66 | 373.24 | 151.28 |
| **Other Main** | 289.29 | 202.00 | 280.49 | 502.49 | 382.00 | 430.54 | 496.96 | 370.00 | 428.47 |
| **Pizza** | 277.23 | 272.99 | 95.25 | 514.50 | 575.50 | 282.90 | 484.06 | 505.41 | 221.19 |
| **Chicken** | 252.42 | 211.40 | 247.10 | 330.99 | 285.00 | 220.31 | 344.46 | 285.00 | 302.93 |
| **Burger** | 209.64 | 211.79 | 97.32 | 348.56 | 332.00 | 201.97 | 353.12 | 332.00 | 192.22 |

**S15 Table.**
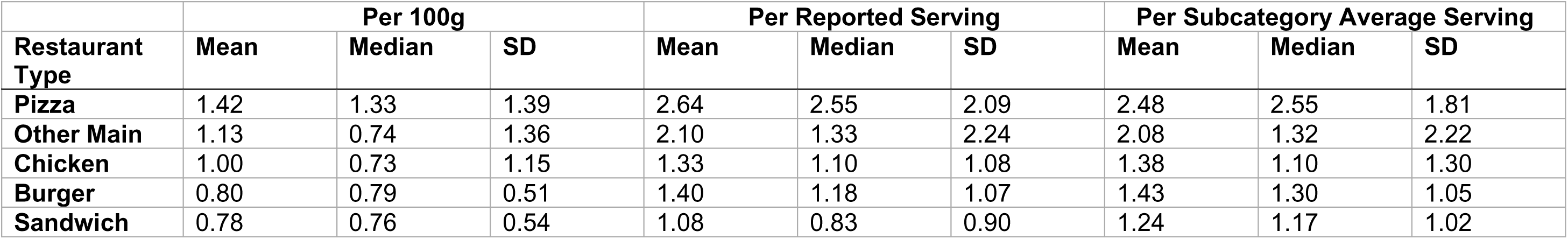
The mean, median, and standard deviation, for Salt per 100g, per recommended serving, and per subcategory average serving, across all menu items in each restaurant group. In descending order by Mean Salt per 100g.

**S16 Table.** The mean, median, and standard deviation, for Sugar per 100g, per recommended serving, and per subcategory average serving, across all menu items in each restaurant group. In descending order by Mean Sugar per 100g.

| Restaurant Type | Per 100g |  |  | Per Reported Serving |  |  | Per Subcategory Average Serving |  |  |
| --- | --- | --- | --- | --- | --- | --- | --- | --- | --- |
|  | Mean | Median | SD | Mean | Median | SD | Mean | Median | SD |
| **Sandwich** | 13.44 | 4.50 | 15.26 | 11.27 | 6.95 | 11.27 | 12.77 | 8.22 | 11.18 |
| **Other Main** | 10.64 | 3.26 | 22.05 | 12.56 | 6.90 | 17.91 | 12.53 | 6.80 | 18.13 |
| **Chicken** | 9.25 | 2.99 | 15.00 | 8.73 | 5.00 | 11.21 | 8.82 | 5.00 | 11.24 |
| **Burger** | 8.25 | 3.59 | 11.61 | 9.53 | 7.30 | 8.99 | 9.58 | 7.42 | 8.86 |
| **Pizza** | 5.83 | 3.80 | 7.44 | 9.25 | 6.50 | 9.67 | 8.54 | 7.34 | 6.43 |

**S17 Table.** The mean, median, and standard deviation, for Fat per 100g, per recommended serving, and per subcategory average serving, across all menu items in each restaurant group. In descending order by Mean Fat per 100g.

| Restaurant Type | Per 100g |  |  | Per Reported Serving |  |  | Per Subcategory Average Serving |  |  |
| --- | --- | --- | --- | --- | --- | --- | --- | --- | --- |
|  | Mean | Median | SD | Mean | Median | SD | Mean | Median | SD |
| Other Main | 14.74 | 9.15 | 18.13 | 24.71 | 18.00 | 23.88 | 24.45 | 17.66 | 24.01 |
| Sandwich | 13.46 | 11.00 | 9.82 | 14.32 | 14.00 | 8.86 | 16.81 | 15.74 | 10.84 |
| Chicken | 11.79 | 9.02 | 13.36 | 14.74 | 13.45 | 11.34 | 15.28 | 13.45 | 14.00 |
| Pizza | 11.56 | 10.20 | 7.69 | 20.02 | 19.22 | 11.88 | 18.83 | 18.22 | 9.61 |
| Burger | 9.17 | 9.47 | 6.41 | 15.50 | 12.50 | 12.39 | 15.64 | 13.00 | 11.54 |

**S18 Table.** The alpha level and p value for pairwise comparisons between restaurants in kcal, sugar, and salt per 100g.

| Outcome Variable | Comparison | Group | Alpha Level (with Bonferroni Correction) | *p* value |
| --- | --- | --- | --- | --- |
| **Kcal per 100g** | Burger King - McDonald's | Burger | 0.050 | 0.003 |
|  | KFC - Nando's | Chicken | 0.050 | 0.061 |
|  | Harvester - Hungry Horse | Other Main | 0.002 | 0.398 |
|  | Harvester - Leon | Other Main | 0.002 | 0.007 |
|  | Harvester - Prezzo | Other Main | 0.002 | 0.120 |
|  | Harvester - Toby Carvery | Other Main | 0.002 | <.001 |
|  | Harvester - Vintage Inns | Other Main | 0.002 | 0.873 |
|  | Harvester - Wagamama | Other Main | 0.002 | <.001 |
|  | Hungry Horse - Leon | Other Main | 0.002 | 0.027 |
|  | Hungry Horse - Prezzo | Other Main | 0.002 | 0.009 |
|  | Hungry Horse - Toby Carvery | Other Main | 0.002 | <.001 |
|  | Hungry Horse - Vintage Inns | Other Main | 0.002 | 0.427 |
|  | Hungry Horse - Wagamama | Other Main | 0.002 | <.001 |
|  | Leon - Prezzo | Other Main | 0.002 | <.001 |
|  | Leon - Toby Carvery | Other Main | 0.002 | 0.434 |
|  | Leon - Vintage Inns | Other Main | 0.002 | 0.015 |
|  | Leon - Wagamama | Other Main | 0.002 | 0.003 |
|  | Prezzo - Toby Carvery | Other Main | 0.002 | <.001 |
|  | Prezzo - Vintage Inns | Other Main | 0.002 | 0.317 |
|  | Prezzo - Wagamama | Other Main | 0.002 | <.001 |
|  | Toby Carvery - Vintage Inns | Other Main | 0.002 | <.001 |
|  | Toby Carvery - Wagamama | Other Main | 0.002 | 0.004 |
|  | Vintage Inns - Wagamama | Other Main | 0.002 | <.001 |
|  | Domino's - Papa John's | Pizza | 0.008 | 0.727 |
|  | Domino's - Pizza Express | Pizza | 0.008 | <.001 |
|  | Domino's - Pizza Hut | Pizza | 0.008 | <.001 |
|  | Papa John's - Pizza Express | Pizza | 0.008 | <.001 |
|  | Papa John's - Pizza Hut | Pizza | 0.008 | <.001 |
|  | Pizza Express - Pizza Hut | Pizza | 0.008 | <.001 |
|  | Caffé Nero - Costa | Sandwich | 0.003 | 0.605 |
|  | Caffé Nero - Greggs | Sandwich | 0.003 | <.001 |
|  | Caffé Nero - Pret | Sandwich | 0.003 | <.001 |
|  | Caffé Nero - Starbucks | Sandwich | 0.003 | 0.001 |
|  | Caffé Nero - Subway | Sandwich | 0.003 | <.001 |
|  | Costa - Greggs | Sandwich | 0.003 | <.001 |
|  | Costa - Pret | Sandwich | 0.003 | <.001 |
|  | Costa - Starbucks | Sandwich | 0.003 | 0.003 |
|  | Costa - Subway | Sandwich | 0.003 | <.001 |
|  | Greggs - Pret | Sandwich | 0.003 | 0.195 |
|  | Greggs - Starbucks | Sandwich | 0.003 | 0.741 |
|  | Greggs - Subway | Sandwich | 0.003 | <.001 |
|  | Pret - Starbucks | Sandwich | 0.003 | 0.230 |
|  | Pret - Subway | Sandwich | 0.003 | 0.001 |
|  | Starbucks - Subway | Sandwich | 0.003 | <.001 |
| **Salt per 100g** | Burger King - McDonald's | Burger | 0.050 | 0.006 |
|  | KFC - Nando's | Chicken | 0.050 | 0.899 |
|  | Harvester - Hungry Horse | Other Main | 0.002 | 0.188 |
|  | Harvester - Leon | Other Main | 0.002 | 0.515 |
|  | Harvester - Prezzo | Other Main | 0.002 | <.001 |
|  | Harvester - Toby Carvery | Other Main | 0.002 | 0.002 |
|  | Harvester - Vintage Inns | Other Main | 0.002 | 0.962 |
|  | Harvester - Wagamama | Other Main | 0.002 | 0.119 |
|  | Hungry Horse - Leon | Other Main | 0.002 | 0.105 |
|  | Hungry Horse - Prezzo | Other Main | 0.002 | <.001 |
|  | Hungry Horse - Toby Carvery | Other Main | 0.002 | <.001 |
|  | Hungry Horse - Vintage Inns | Other Main | 0.002 | 0.355 |
|  | Hungry Horse - Wagamama | Other Main | 0.002 | 0.006 |
|  | Leon - Prezzo | Other Main | 0.002 | <.001 |
|  | Leon - Toby Carvery | Other Main | 0.002 | 0.103 |
|  | Leon - Vintage Inns | Other Main | 0.002 | 0.553 |
|  | Leon - Wagamama | Other Main | 0.002 | 0.527 |
|  | Prezzo - Toby Carvery | Other Main | 0.002 | <.001 |
|  | Prezzo - Vintage Inns | Other Main | 0.002 | <.001 |
|  | Prezzo - Wagamama | Other Main | 0.002 | <.001 |
|  | Toby Carvery - Vintage Inns | Other Main | 0.002 | 0.015 |
|  | Toby Carvery - Wagamama | Other Main | 0.002 | 0.278 |
|  | Vintage Inns - Wagamama | Other Main | 0.002 | 0.191 |
|  | Domino's - Papa John's | Pizza | 0.008 | 0.903 |
|  | Domino's - Pizza Express | Pizza | 0.008 | <.001 |
|  | Domino's - Pizza Hut | Pizza | 0.008 | 0.013 |
|  | Papa John's - Pizza Express | Pizza | 0.008 | <.001 |
|  | Papa John's - Pizza Hut | Pizza | 0.008 | 0.016 |
|  | Pizza Express - Pizza Hut | Pizza | 0.008 | <.001 |
|  | Caffé Nero - Costa | Sandwich | 0.003 | 0.968 |
|  | Caffé Nero - Greggs | Sandwich | 0.003 | <.001 |
|  | Caffé Nero - Pret | Sandwich | 0.003 | 0.009 |
|  | Caffé Nero - Starbucks | Sandwich | 0.003 | 0.499 |
|  | Caffé Nero - Subway | Sandwich | 0.003 | <.001 |
|  | Costa - Greggs | Sandwich | 0.003 | <.001 |
|  | Costa - Pret | Sandwich | 0.003 | 0.008 |
|  | Costa - Starbucks | Sandwich | 0.003 | 0.472 |
|  | Costa - Subway | Sandwich | 0.003 | <.001 |
|  | Greggs - Pret | Sandwich | 0.003 | 0.139 |
|  | Greggs - Starbucks | Sandwich | 0.003 | <.001 |
|  | Greggs - Subway | Sandwich | 0.003 | 0.551 |
|  | Pret - Starbucks | Sandwich | 0.003 | 0.007 |
|  | Pret - Subway | Sandwich | 0.003 | 0.076 |
|  | Starbucks - Subway | Sandwich | 0.003 | <.001 |
| **Sugar per 100g** | Burger King - McDonald's | Burger | 0.050 | 0.418 |
|  | KFC - Nando's | Chicken | 0.050 | 0.235 |
|  | Harvester - Hungry Horse | Other Main | 0.002 | <.001 |
|  | Harvester - Leon | Other Main | 0.002 | 0.025 |
|  | Harvester - Prezzo | Other Main | 0.002 | <.001 |
|  | Harvester - Toby Carvery | Other Main | 0.002 | 0.167 |
|  | Harvester - Vintage Inns | Other Main | 0.002 | 0.072 |
|  | Harvester - Wagamama | Other Main | 0.002 | 0.010 |
|  | Hungry Horse - Leon | Other Main | 0.002 | 0.824 |
|  | Hungry Horse - Prezzo | Other Main | 0.002 | 0.698 |
|  | Hungry Horse - Toby Carvery | Other Main | 0.002 | 0.023 |
|  | Hungry Horse - Vintage Inns | Other Main | 0.002 | 0.443 |
|  | Hungry Horse - Wagamama | Other Main | 0.002 | 0.774 |
|  | Leon - Prezzo | Other Main | 0.002 | 0.633 |
|  | Leon - Toby Carvery | Other Main | 0.002 | 0.195 |
|  | Leon - Vintage Inns | Other Main | 0.002 | 0.672 |
|  | Leon - Wagamama | Other Main | 0.002 | 0.985 |
|  | Prezzo - Toby Carvery | Other Main | 0.002 | 0.010 |
|  | Prezzo - Vintage Inns | Other Main | 0.002 | 0.307 |
|  | Prezzo - Wagamama | Other Main | 0.002 | 0.558 |
|  | Toby Carvery - Vintage Inns | Other Main | 0.002 | 0.417 |
|  | Toby Carvery - Wagamama | Other Main | 0.002 | 0.134 |
|  | Vintage Inns - Wagamama | Other Main | 0.002 | 0.652 |
|  | Domino's - Papa John's | Pizza | 0.008 | 0.469 |
|  | Domino's - Pizza Express | Pizza | 0.008 | <.001 |
|  | Domino's - Pizza Hut | Pizza | 0.008 | <.001 |
|  | Papa John's - Pizza Express | Pizza | 0.008 | <.001 |
|  | Papa John's - Pizza Hut | Pizza | 0.008 | 0.001 |
|  | Pizza Express - Pizza Hut | Pizza | 0.008 | 0.138 |
|  | Caffé Nero - Costa | Sandwich | 0.003 | 0.588 |
|  | Caffé Nero - Greggs | Sandwich | 0.003 | <.001 |
|  | Caffé Nero - Pret | Sandwich | 0.003 | <.001 |
|  | Caffé Nero - Starbucks | Sandwich | 0.003 | 0.525 |
|  | Caffé Nero - Subway | Sandwich | 0.003 | <.001 |
|  | Costa - Greggs | Sandwich | 0.003 | <.001 |
|  | Costa - Pret | Sandwich | 0.003 | <.001 |
|  | Costa - Starbucks | Sandwich | 0.003 | 0.284 |
|  | Costa - Subway | Sandwich | 0.003 | <.001 |
|  | Greggs - Pret | Sandwich | 0.003 | 0.258 |
|  | Greggs - Starbucks | Sandwich | 0.003 | 0.004 |
|  | Greggs - Subway | Sandwich | 0.003 | 0.006 |
|  | Pret - Starbucks | Sandwich | 0.003 | <.001 |
|  | Pret - Subway | Sandwich | 0.003 | 0.075 |
|  | Starbucks - Subway | Sandwich | 0.003 | <.001 |

**S19 Table.** The proportion of menu items meeting sugar, salt, calorie, and all applicable targets, for each restaurant group. In descending order by proportion of menu items meeting all applicable targets.

|  | Calorie Targets |  |  | Salt Targets |  |  | Sugar Targets |  |  | All Applicable Targets |  |  |
| --- | --- | --- | --- | --- | --- | --- | --- | --- | --- | --- | --- | --- |
| Restaura<br>nt Type | No. of<br>eligib<br>le<br>menu<br>items | Proporti<br>on of<br>menu<br>items<br>meeting<br>target<br>(%) | Range in<br>proportio<br>n across<br>restaura<br>nts (%) | No. of<br>eligib<br>le<br>menu<br>items | Proporti<br>on of<br>menu<br>items<br>meeting<br>target<br>(%) | Range in<br>proportio<br>n across<br>restaura<br>nts (%) | No. of<br>eligib<br>le<br>menu<br>items | Proporti<br>on of<br>menu<br>items<br>meeting<br>target<br>(%) | Range in<br>proportio<br>n across<br>restaura<br>nts (%) | No. of<br>eligib<br>le<br>menu<br>items | Proporti<br>on of<br>menu<br>items<br>meeting<br>target<br>(%) | Range in<br>proportio<br>n across<br>restaura<br>nts (%) |
| **Burger** | 102 | 69 | 41 | 115 | 80 | 10 | 36 | 53 | 59 | 149 | 59 | 33 |
| **Chicken** | 124 | 78 | 19 | 116 | 66 | 6 | 23 | 0 | 0 | 166 | 58 | 5 |
| **Sandwic**<br>**h** | 369 | 72 | 20 | 566 | 69 | 33 | 257 | 35 | 34 | 657 | 48 | 46 |
| **Other**<br>**Main** | 707 | 59 | 42 | 715 | 53 | 53 | 197 | 42 | 62 | 1018 | 46 | 37 |
| **Pizza** | 846 | 53 | 26 | 832 | 49 | 59 | 65 | 23 | 19 | 961 | 32 | 33 |

**S20 Table.**
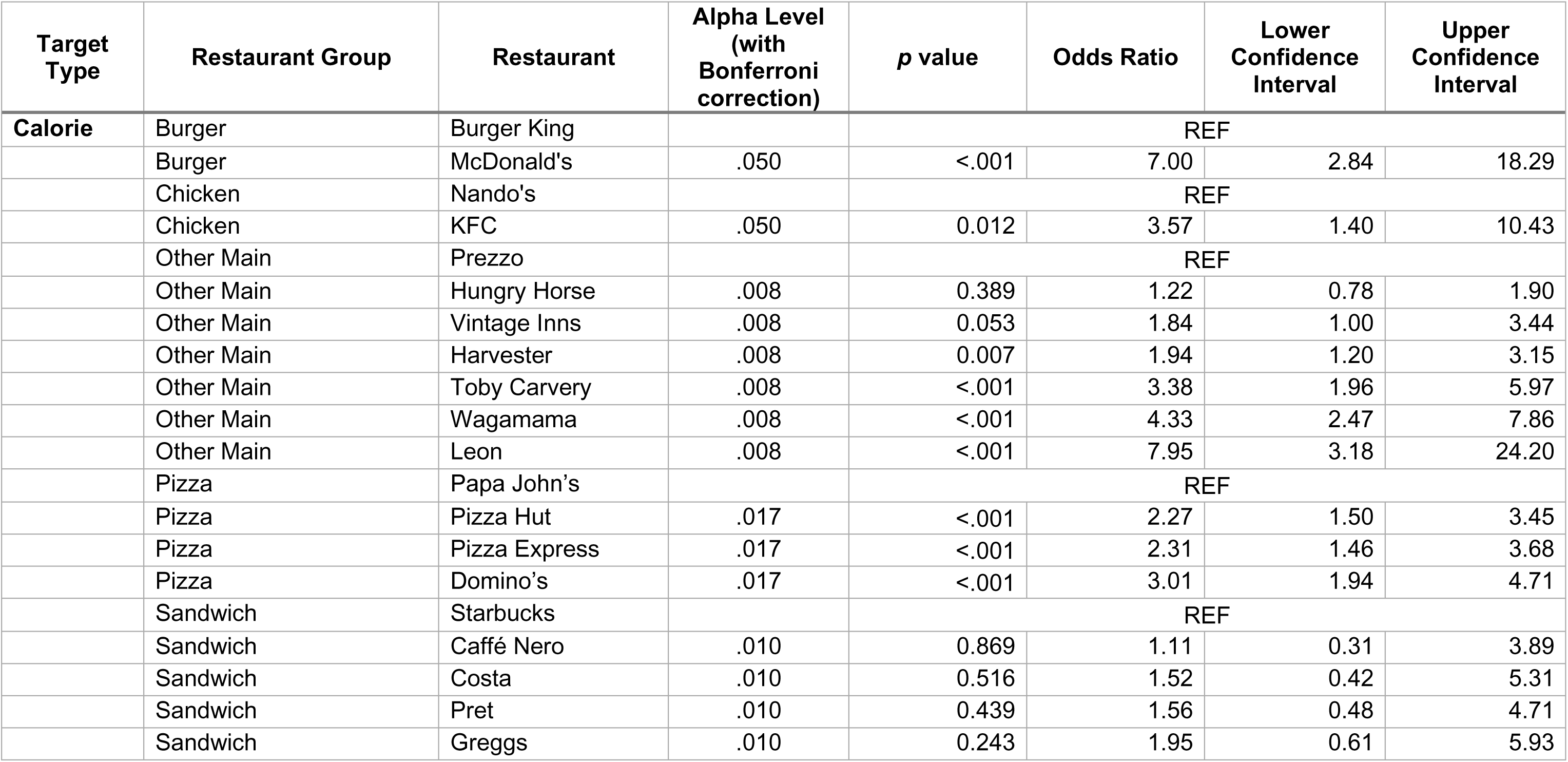
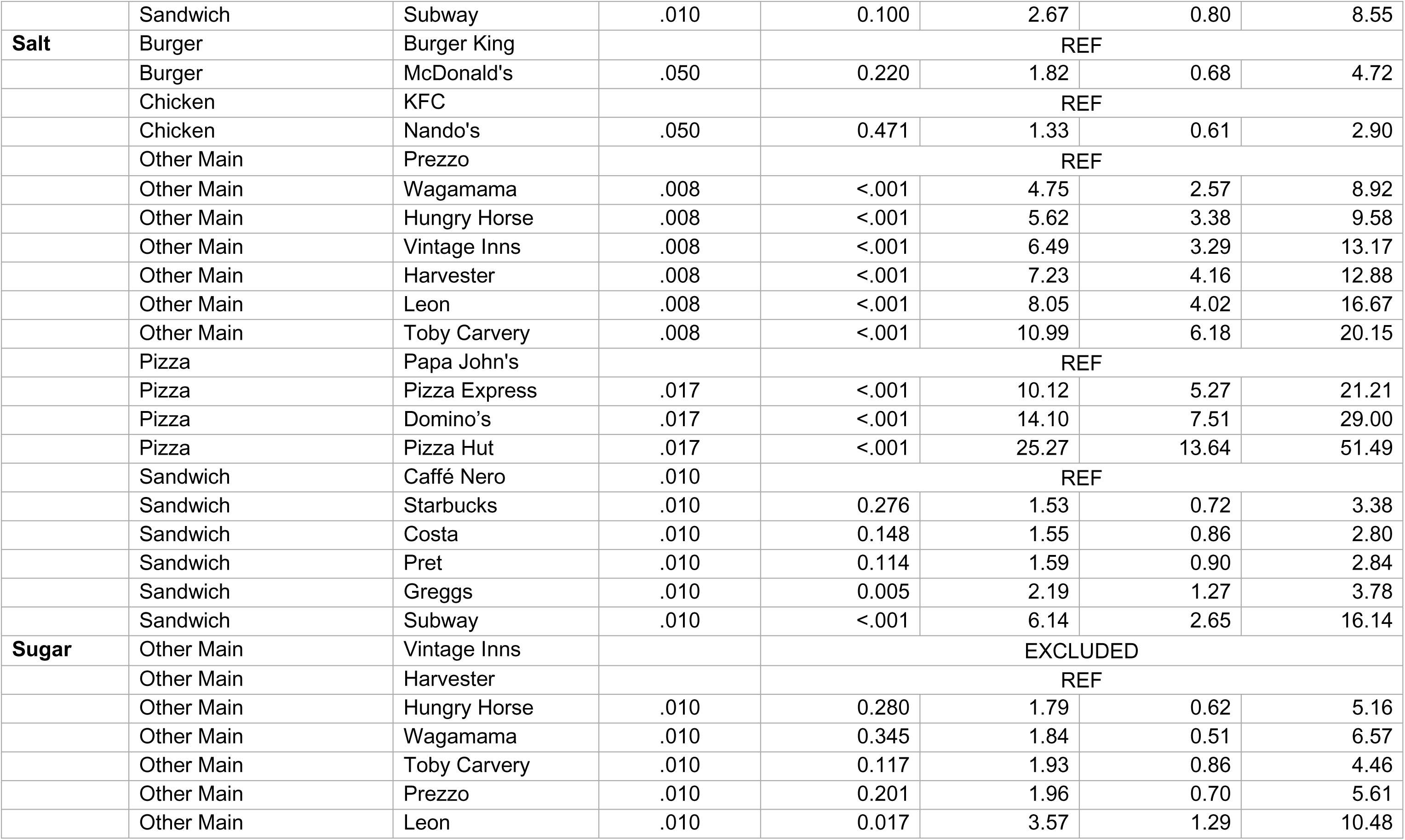
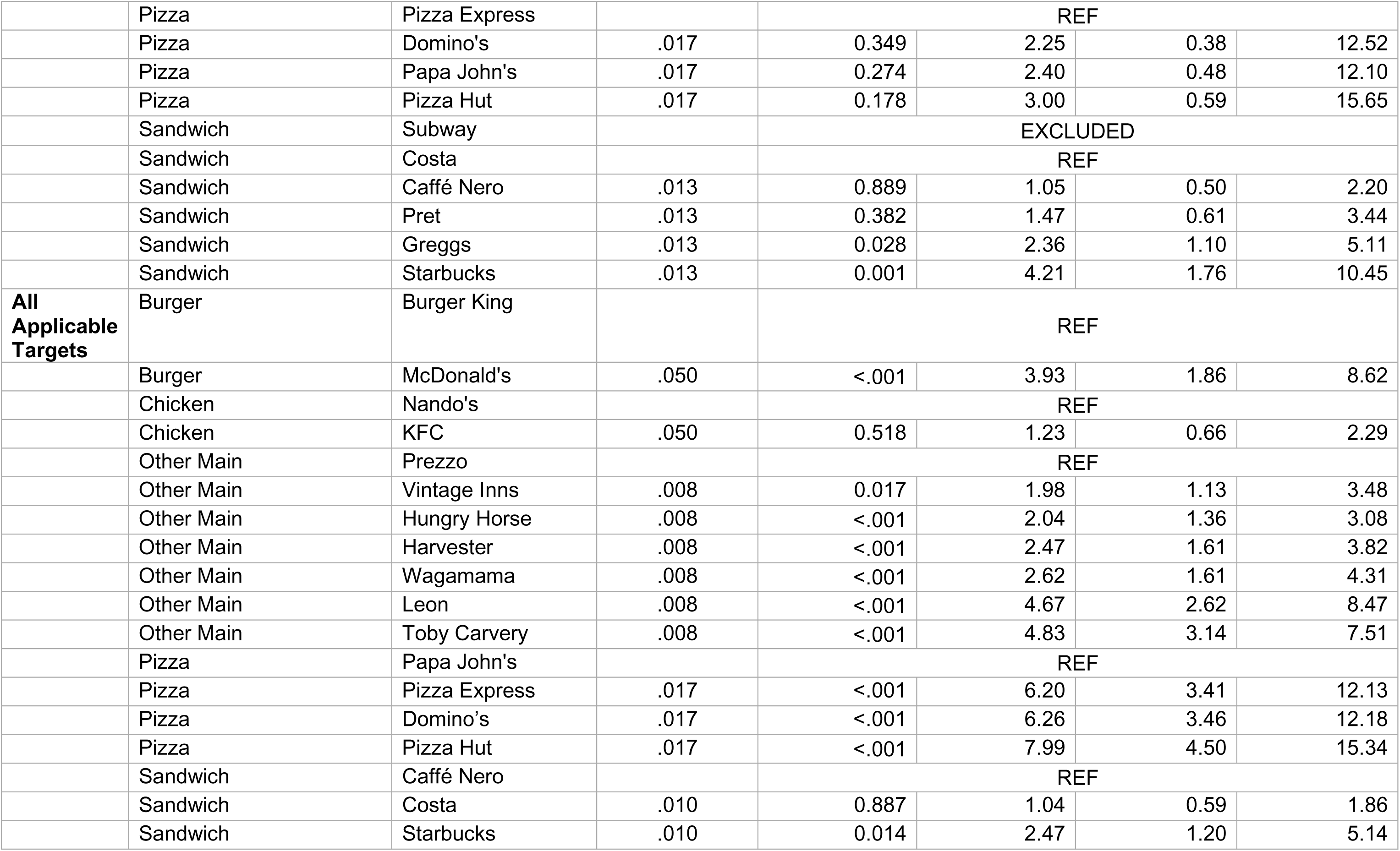
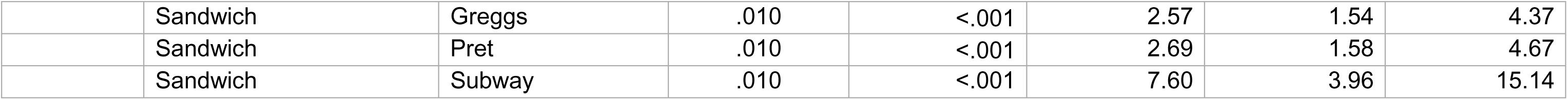
The p value, odds ratio, and confidence intervals, for logistic regressions models with the predictor of restaurant and outcome of target met (yes or no) for calorie, salt, sugar, and all applicable targets. ‘REF’ indicates the reference group for each model, being the restaurant with the lowest adherence to that target within their group.

| Target Type | Restaurant Group | Restaurant | Alpha Level (with Bonferroni correction) | p value | Odds Ratio | Lower Confidence Interval | Upper Confidence Interval |
| --- | --- | --- | --- | --- | --- | --- | --- |
| Calorie | Burger | Burger King |  | REF |  |  |  |
|  | Burger | McDonald's | .050 | <.001 | 7.00 | 2.84 | 18.29 |
|  | Chicken | Nando's |  | REF |  |  |  |
|  | Chicken | KFC | .050 | 0.012 | 3.57 | 1.40 | 10.43 |
|  | Other Main | Prezzo |  | REF |  |  |  |
|  | Other Main | Hungry Horse | .008 | 0.389 | 1.22 | 0.78 | 1.90 |
|  | Other Main | Vintage Inns | .008 | 0.053 | 1.84 | 1.00 | 3.44 |
|  | Other Main | Harvester | .008 | 0.007 | 1.94 | 1.20 | 3.15 |
|  | Other Main | Toby Carvery | .008 | <.001 | 3.38 | 1.96 | 5.97 |
|  | Other Main | Wagamama | .008 | <.001 | 4.33 | 2.47 | 7.86 |
|  | Other Main | Leon | .008 | <.001 | 7.95 | 3.18 | 24.20 |
|  | Pizza | Papa John's |  | REF |  |  |  |
|  | Pizza | Pizza Hut | .017 | <.001 | 2.27 | 1.50 | 3.45 |
|  | Pizza | Pizza Express | .017 | <.001 | 2.31 | 1.46 | 3.68 |
|  | Pizza | Domino's | .017 | <.001 | 3.01 | 1.94 | 4.71 |
|  | Sandwich | Starbucks |  | REF |  |  |  |
|  | Sandwich | Caffé Nero | .010 | 0.869 | 1.11 | 0.31 | 3.89 |
|  | Sandwich | Costa | .010 | 0.516 | 1.52 | 0.42 | 5.31 |
|  | Sandwich | Pret | .010 | 0.439 | 1.56 | 0.48 | 4.71 |
|  | Sandwich | Greggs | .010 | 0.243 | 1.95 | 0.61 | 5.93 |
|  | Sandwich | Subway | .010 | 0.100 | 2.67 | 0.80 | 8.55 |
| **Salt** | Burger | Burger King |  | REF |  |  |  |
|  | Burger | McDonald's | .050 | 0.220 | 1.82 | 0.68 | 4.72 |
|  | Chicken | KFC |  | REF |  |  |  |
|  | Chicken | Nando's | .050 | 0.471 | 1.33 | 0.61 | 2.90 |
|  | Other Main | Prezzo |  | REF |  |  |  |
|  | Other Main | Wagamama | .008 | <.001 | 4.75 | 2.57 | 8.92 |
|  | Other Main | Hungry Horse | .008 | <.001 | 5.62 | 3.38 | 9.58 |
|  | Other Main | Vintage Inns | .008 | <.001 | 6.49 | 3.29 | 13.17 |
|  | Other Main | Harvester | .008 | <.001 | 7.23 | 4.16 | 12.88 |
|  | Other Main | Leon | .008 | <.001 | 8.05 | 4.02 | 16.67 |
|  | Other Main | Toby Carvery | .008 | <.001 | 10.99 | 6.18 | 20.15 |
|  | Pizza | Papa John's |  | REF |  |  |  |
|  | Pizza | Pizza Express | .017 | <.001 | 10.12 | 5.27 | 21.21 |
|  | Pizza | Domino's | .017 | <.001 | 14.10 | 7.51 | 29.00 |
|  | Pizza | Pizza Hut | .017 | <.001 | 25.27 | 13.64 | 51.49 |
|  | Sandwich | Caffé Nero | .010 | REF |  |  |  |
|  | Sandwich | Starbucks | .010 | 0.276 | 1.53 | 0.72 | 3.38 |
|  | Sandwich | Costa | .010 | 0.148 | 1.55 | 0.86 | 2.80 |
|  | Sandwich | Pret | .010 | 0.114 | 1.59 | 0.90 | 2.84 |
|  | Sandwich | Greggs | .010 | 0.005 | 2.19 | 1.27 | 3.78 |
|  | Sandwich | Subway | .010 | <.001 | 6.14 | 2.65 | 16.14 |
| **Sugar** | Other Main | Vintage Inns |  | EXCLUDED |  |  |  |
|  | Other Main | Harvester |  | REF |  |  |  |
|  | Other Main | Hungry Horse | .010 | 0.280 | 1.79 | 0.62 | 5.16 |
|  | Other Main | Wagamama | .010 | 0.345 | 1.84 | 0.51 | 6.57 |
|  | Other Main | Toby Carvery | .010 | 0.117 | 1.93 | 0.86 | 4.46 |
|  | Other Main | Prezzo | .010 | 0.201 | 1.96 | 0.70 | 5.61 |
|  | Other Main | Leon | .010 | 0.017 | 3.57 | 1.29 | 10.48 |
|  | Pizza | Pizza Express |  | REF |  |  |  |
|  | Pizza | Domino's | .017 | 0.349 | 2.25 | 0.38 | 12.52 |
|  | Pizza | Papa John's | .017 | 0.274 | 2.40 | 0.48 | 12.10 |
|  | Pizza | Pizza Hut | .017 | 0.178 | 3.00 | 0.59 | 15.65 |
|  | Sandwich | Subway |  | EXCLUDED |  |  |  |
|  | Sandwich | Costa |  | REF |  |  |  |
|  | Sandwich | Caffé Nero | .013 | 0.889 | 1.05 | 0.50 | 2.20 |
|  | Sandwich | Pret | .013 | 0.382 | 1.47 | 0.61 | 3.44 |
|  | Sandwich | Greggs | .013 | 0.028 | 2.36 | 1.10 | 5.11 |
|  | Sandwich | Starbucks | .013 | 0.001 | 4.21 | 1.76 | 10.45 |
| **All**<br>**Applicable**<br>**Targets** | Burger | Burger King |  | REF |  |  |  |
|  | Burger | McDonald's | .050 | <.001 | 3.93 | 1.86 | 8.62 |
|  | Chicken | Nando's |  | REF |  |  |  |
|  | Chicken | KFC | .050 | 0.518 | 1.23 | 0.66 | 2.29 |
|  | Other Main | Prezzo |  | REF |  |  |  |
|  | Other Main | Vintage Inns | .008 | 0.017 | 1.98 | 1.13 | 3.48 |
|  | Other Main | Hungry Horse | .008 | <.001 | 2.04 | 1.36 | 3.08 |
|  | Other Main | Harvester | .008 | <.001 | 2.47 | 1.61 | 3.82 |
|  | Other Main | Wagamama | .008 | <.001 | 2.62 | 1.61 | 4.31 |
|  | Other Main | Leon | .008 | <.001 | 4.67 | 2.62 | 8.47 |
|  | Other Main | Toby Carvery | .008 | <.001 | 4.83 | 3.14 | 7.51 |
|  | Pizza | Papa John's |  | REF |  |  |  |
|  | Pizza | Pizza Express | .017 | <.001 | 6.20 | 3.41 | 12.13 |
|  | Pizza | Domino's | .017 | <.001 | 6.26 | 3.46 | 12.18 |
|  | Pizza | Pizza Hut | .017 | <.001 | 7.99 | 4.50 | 15.34 |
|  | Sandwich | Caffé Nero |  | REF |  |  |  |
|  | Sandwich | Costa | .010 | 0.887 | 1.04 | 0.59 | 1.86 |
|  | Sandwich | Starbucks | .010 | 0.014 | 2.47 | 1.20 | 5.14 |
|  | Sandwich | Greggs | .010 | <.001 | 2.57 | 1.54 | 4.37 |
|  | Sandwich | Pret | .010 | <.001 | 2.69 | 1.58 | 4.67 |
|  | Sandwich | Subway | .010 | <.001 | 7.60 | 3.96 | 15.14 |

**S21 Table.** The alpha level and p value for pairwise comparisons between restaurants in kcal, sugar, and salt per 100g, for the sensitivity analysis.

| Outcome Variable | Comparison | Group | Alpha Level (with Bonferroni Correction) | p value |
| --- | --- | --- | --- | --- |
| **Kcal per 100g** | Burger | McDonald's - Burger King | 0.050 | 0.001 |
|  | Chicken | KFC – Nando's | 0.050 | 0.288 |
|  | Sandwich | Pret - Caffé Nero | 0.003 | <.001 |
|  | Sandwich | Pret - Costa | 0.003 | <.001 |
|  | Sandwich | Pret - Greggs | 0.003 | 0.406 |
|  | Sandwich | Pret - Starbucks | 0.003 | 0.372 |
|  | Sandwich | Pret - Subway | 0.003 | <.001 |
| **Salt per 100g** | Burger | McDonald's - Burger King | 0.050 | 0.009 |
|  | Chicken | KFC – Nando's | 0.050 | 0.877 |
|  | Sandwich | Pret - Caffé Nero | 0.003 | 0.010 |
|  | Sandwich | Pret - Costa | 0.003 | 0.009 |
|  | Sandwich | Pret - Greggs | 0.003 | 0.162 |
|  | Sandwich | Pret - Starbucks | 0.003 | 0.007 |
|  | Sandwich | Pret - Subway | 0.003 | 0.090 |
| **Sugar per 100g** | Burger | McDonald's - Burger King | 0.050 | 0.383 |
|  | Chicken | KFC – Nando's | 0.050 | 0.012 |
|  | Sandwich | Pret - Caffé Nero | 0.003 | <.001 |
|  | Sandwich | Pret - Costa | 0.003 | <.001 |
|  | Sandwich | Pret - Greggs | 0.003 | 0.257 |
|  | Sandwich | Pret - Starbucks | 0.003 | <.001 |
|  | Sandwich | Pret - Subway | 0.003 | 0.082 |

**S22 Table.** The p value, odds ratio, and confidence intervals, for the logistic regression models conducted for the sensitivity analysis.

| Target Type | Restaurant Group | Restaurant | Alpha Level (with Bonferroni correction) | p value | Odds Ratio | Lower Confidence Interval | Upper Confidence Interval |
| --- | --- | --- | --- | --- | --- | --- | --- |
| Calorie | Burger | Burger King |  | REF |  |  |  |
|  | Burger | McDonald's | .050 | <.001 | 6.46 | 2.54 | 17.46 |
|  | Chicken | Nando's |  | REF |  |  |  |
|  | Chicken | KFC | .050 | 0.078 | 2.48 | 0.95 | 7.34 |
|  | Sandwich | Starbucks |  | REF |  |  |  |
|  | Sandwich | Caffé Nero | .010 | 0.869 | 1.11 | 0.31 | 3.89 |
|  | Sandwich | Costa | .010 | 0.516 | 1.52 | 0.42 | 5.31 |
|  | Sandwich | Pret | .010 | 0.516 | 1.45 | 0.45 | 4.43 |
|  | Sandwich | Greggs | .010 | 0.243 | 1.95 | 0.61 | 5.93 |
|  | Sandwich | Subway | .010 | 0.100 | 2.67 | 0.80 | 8.55 |
| Salt | Burger | Burger King |  | REF |  |  |  |
|  | Burger | McDonald's | .050 | 0.279 | 1.74 | 0.62 | 4.69 |
|  | Chicken | KFC |  | REF |  |  |  |
|  | Chicken | Nando's | .050 | 0.082 | 2.07 | 0.92 | 4.76 |
|  | Sandwich | Caffé Nero |  | REF |  |  |  |
|  | Sandwich | Starbucks | .010 | 0.276 | 1.53 | 0.72 | 3.38 |
|  | Sandwich | Costa | .010 | 0.148 | 1.55 | 0.86 | 2.80 |
|  | Sandwich | Pret | .010 | 0.187 | 1.48 | 0.83 | 2.67 |
|  | Sandwich | Greggs | .010 | 0.005 | 2.19 | 1.27 | 3.78 |
|  | Sandwich | Subway | .010 | <.001 | 6.14 | 2.65 | 16.14 |
| Sugar Targets | Sandwich | Subway |  | EXCLUDED |  |  |  |
|  | Sandwich | Costa |  | REF |  |  |  |
|  | Sandwich | Caffé Nero | .013 | 0.889 | 1.05 | 0.50 | 2.20 |
|  | Sandwich | Pret | .013 | 0.508 | 1.34 | 0.55 | 3.19 |
|  | Sandwich | Greggs | .013 | 0.028 | 2.36 | 1.10 | 5.11 |
|  | Sandwich | Starbucks | .013 | 0.001 | 4.21 | 1.76 | 10.45 |
| **All Applicable Targets** | Burger | Burger King |  | REF |  |  |  |
|  | Burger | McDonald's | .050 | <.001 | 3.87 | 1.79 | 8.65 |
|  | Chicken | Nando's |  | REF |  |  |  |
|  | Chicken | KFC | .050 | 0.708 | 1.13 | 0.59 | 2.18 |
|  | Sandwich | Caffé Nero |  | REF |  |  |  |
|  | Sandwich | Costa | .010 | 0.887 | 1.04 | 0.59 | 1.86 |
|  | Sandwich | Starbucks | .010 | 0.014 | 2.47 | 1.20 | 5.14 |
|  | Sandwich | Greggs | .010 | <.001 | 2.57 | 1.54 | 4.37 |
|  | Sandwich | Pret | .010 | 0.001 | 2.48 | 1.44 | 4.34 |
|  | Sandwich | Subway | .010 | <.001 | 7.60 | 3.96 | 15.14 |

